# Development of an ensemble machine learning prognostic model to predict 60-day risk of major adverse cardiac events in adults with chest pain

**DOI:** 10.1101/2021.03.08.21252615

**Authors:** Chris J. Kennedy, Dustin G. Mark, Jie Huang, Mark J. van der Laan, Alan E. Hubbard, Mary E. Reed

**Affiliations:** Kaiser Permanente Northern California, Oakland, CA, USA; Department of Biomedical Informatics, Harvard Medical School, Boston, MA, USA; Department of Surgery, Beth Israel Deaconess Medical Center, Boston, MA, USA; Division of Biostatistics, UC Berkeley, Berkeley, CA, USA

## Abstract

**Background:** Chest pain is the second leading reason for emergency department (ED) visits and is commonly identified as a leading driver of low-value health care. Accurate identification of patients at low risk of major adverse cardiac events (MACE) is important to improve resource allocation and reduce over-treatment.

**Objectives:** We assessed machine learning (ML) methods and electronic health record (EHR) covariate collection for MACE prediction. We aimed to maximize the pool of low-risk patients that were accurately predicted to have less than 0.5% MACE risk and could be eligible for reduced testing (“rule-out” strategy).

**Population Studied:** 116,764 adult patients presenting with chest pain in the ED between 2013 and 2015 and evaluated for potential acute coronary syndrome (ACS). 60-day MACE rate was 2%.

**Setting:** 21 emergency departments within the Kaiser Permanente Northern California integrated health system. Data analysis was performed May 2018 to August 2021.

**Methods:** We evaluated ML algorithms (lasso, splines, random forest, extreme gradient boosting, Bayesian additive regression trees) and SuperLearner stacked ensembling. We tuned ML hyperparameters through nested ensembling, and imputed missing values with generalized low-rank models (GLRM). Performance was benchmarked against individual biomarkers, validated clinical risk scores, decision trees, and logistic regression. We assessed clinical utility through net benefit analysis and explained the models through variable importance ranking and accumulated local effect plots.

**Results:** The SuperLearner ensemble provided the best cross-validated discrimination with areas under the curve of 0.15 for precision-recall (PR-AUC) and 0.87 for receiver operating characteristic (ROC-AUC), and the best accuracy with an index of prediction accuracy of 0.07. The ensemble’s risk estimates were miscalibrated by 0.2 percentage points on average, and dominated the net benefit analysis at all examined thresholds. At a 0.5% threshold the ensemble model yielded 32 benefit-adjusted workups avoided per 100 patients, compared to 25 for logistic regression and 2-14 for clinical risk scores. The most important predictors were age, troponin, clinical risk scores, and electrocardiogram. GLRM achieved a 90% average reduction in reconstruction error compared to median-mode imputation.

**Conclusion:** Combining ML algorithms with a broad set of EHR covariates improved MACE risk prediction and would reduce over-treatment compared to simpler alternatives, while providing calibrated predictions and interpretability. Patients should receive targeted benefit in their care from thorough detection of nuanced health patterns via ML.

The omission of prediction from the major goals of basic medical science has impoverished the intellectual content of clinical work, since a modern clinician’s main challenge in the care of patients is to make predictions.
Alvan Feinstein, 1983

## 1 Introduction

Chest pain is the second leading reason for emergency department visits (Rui et al. 2016) and is commonly identified as a leading driver of low-value health care (Sabbatini et al. 2014). Workup protocols in patients with chest pain are designed to diagnose the potential for major adverse cardiac events (MACE) (Kontos et al. 2010). Missed diagnoses of MACE can be cause for medico-legal action, which may encourage conservative testing without health benefit. Accurate identification of patients at low risk of MACE is important to improve resource allocation and reduce overtreatment (Amsterdam et al. 2010). Risk scores aim to identify patients eligible for early discharge (i.e. a “rule-out” strategy), avoiding additional stress testing and cardiac imaging that is unlikely to be of benefit (Greenslade et al. 2018). The primary biomarkers used for initial triage are elevated cardiac troponin, a sensitive marker of cardiac injury measured serially, and repeated electrocardiograms (Scirica 2010; Smith et al. 2015).

Previous work has focused on the development and validation of additive risk scores as decision aids for risk stratification. Such scores examine a small number of biomarkers and demographics, summarize those predictors into qualitative, ordinal levels, and use a weighted sum to allocate patients into risk categories. Standard risk scores are HEART (History, ECG, Age, Risk factors and Troponin) and EDACS (Emergency Department Assessment of Chest Pain Score - M. Than, Flaws, et al. 2014). HEART is most commonly used in North America, although EDACS has similar performance characteristics (Mark, J. Huang, U. Chettipally, et al. 2018). Effective risk scores will stratify patients across risk levels such that the qualitative “low risk” group will have sufficiently low risk of short-term MACE that those patients can be discharged without additional workup. An ineffective risk score would underestimate the risk in the “low risk” group and lead to an optimistic early discharge policy that misses future MACE.

### 1.1 Objectives

Building on recent work (Mark, J. Huang, U. Chettipally, et al. 2018; Mark, J. Huang, Kene, et al. 2021), we sought to assess the performance of machine learning (ML) methods at predicting MACE among emergency department patients with chest pain. We hypothesized that ML could improve upon existing validated risk scores through a more thorough integration of predictors. Our clinical objective was to maximize the pool of low-risk patients accurately predicted to have less than 0.5% MACE risk and eligible for reduced testing. We highlight the use of net benefit analysis to achieve this goal. The primary threshold of 0.5% risk has previously been identified as an acceptable risk by a majority of emergency physicians for early discharge (M. Than, Herbert, et al. 2013). We also examined secondary thresholds of 1.0% and 2.0% given varying risk tolerance thresholds among practicing emergency physicians (ibid.), as well as variation in patient risk preferences.

A reasonable assessment of ML performance could only be made in comparison to realistic alternative options. We compared ML performance to simpler indicators of risk: key biomarkers (troponin, electrocardiogram), validated clinical risk scores (History, ECG, Age, Risk factors and Troponin [HEART] and Emergency Department Assessment of Chest pain Score [EDACS]), as well as simpler models: decision trees, logistic regression, and ordinary least squares (OLS). If simpler algorithms remain preferred, the ML results can at least approximate the best achievable performance, and so serve as benchmark standards when considering more restrictive algorithms.

Should machine learning successfully reduce unnecessary cardiac workups compared to benchmarks, its next hurdle for adoption is interpretability. Clinicians may be willing to forgo maximum predictive accuracy for the sake of understanding how individual predictors influence the output of the algorithm. Through analytical effort it may be possible to provide sufficient interpretability for clinicians to accept the complication of machine learning and the benefit of the (potentially) improved predictive accuracy.

## 2 Data and Methods

### 2.1 Source of data

Our study was sourced from the electronic health record (EHR) of 21 emergency departments (EDs) within Kaiser Permanente Northern California, an integrated health care delivery system with over 1 million annual ED visits.

### 2.2 Participants

All adult patients were retrospectively included if they had received cardiac troponin testing in the emergency department between 2013 and 2015 and either presented with a chief complaint of chest pain or chest discomfort, or whose ED physician had assigned them a primary or secondary ICD-coded diagnosis of chest pain. The latter inclusion criterion is important because patients may complain of “anginal equivalents” (such as shortness of breath) in lieu of overt chest pain (Amsterdam et al. 2010). The initial inclusion pool had a 60-day MACE rate of 8.0%. Patients were excluded if they had a MACE diagnosis in the ED or within 30 days prior to ED visit, alternative non-ACS diagnoses at index visit (e.g. pneumonia, pneumothorax, or traumatic injury), could not be tracked due to lack of active health plan membership during the study (except in cases of death), or had a troponin I > 99th percentile upper limit of normal given the dominant predictive value of elevated troponin values for adverse outcomes in both patients with acute coronary syndromes and in the general population (Bonaca et al. 2010; De Lemos et al. 2010). Patients were excluded if their smoking status was unknown, a key marker of low-quality data. The final study cohort consisted of 116,764 patients with a 60-day MACE incidence of 1.88%. A fourth-generation troponin assay was used during the study period (AccuTnI+3, Beckman-Couleter, Brea, CA, USA).

### 2.3 Outcome

Our outcome was MACE incidence within 60 days of the index visit. We defined MACE as myocardial infarction, cardiogenic shock, cardiac arrest, or death.

### 2.4 Predictors

We extracted 74 predictors from the EHR, including vitals, labs, history, demographics, qualitative interpretation of ECG imaging, regular expression-based features from clinical notes, and missingness indicators. Details are provided in eTable 1.

### 2.5 Missing data

We created missingness indicators for each predictor with missing values (see eTable 1). Inclusion of missingness indicators often improves predictive performance (Agor et al. 2019; Sperrin et al. 2020), in part because it can reflect the information-seeking behavior of clinicians stemming from medical diagnosis and evaluation (Agniel et al. 2018; Groenwold 2020; Sisk et al. 2021). The set of missingness indicators was analyzed for perfect collinearity, and duplicate indicators were dropped.

Missing predictor values were imputed by factorizing the raw data matrix with generalized low-rank models (GLRM) (Schuler et al. 2016; Udell et al. 2016), a generalization of principal component analysis and matrix completion methods designed for mixed type data frames (continuous, categorical, ordinal, or binary variables). GLRM decomposes the predictor data into a matrix of components and matrix of archetypes, including possible L1 or L2 penalization. Multiplying these two matrices reconstructs the original data frame, imputing any data entries with missing values; it will also allow imputation of missing values in future observations, even for predictors with no missingness in the current dataset. Additional details are provided in the supplemental information.

We evaluated the benefit of the GLRM imputation by comparing imputed to known values, among variables with missingness. The root mean-squared reconstruction error metric was calculated for each variable and imputation method. We could then estimate the percentage improvement in reconstruction error for the GLRM imputation for each variable compared to median/mode imputation.

GLRM imputation greatly increased the number of unique values (cardinality) for continuous variables, which would have a negative performance impact on tree-based algorithms that test every unique value for a potential split. To avoid that performance drop, we compressed high-cardinality imputed predictors to 200 unique values using penalized histograms (Rozenholc et al. 2010).

### 2.6 Prediction algorithms

Our prediction algorithm selection strategy focused on well-known algorithms that have shown strong performance in prior research, including both linear and tree-based estimation. The tree-based algorithms were random forest [RF] (Breiman 2001), extreme gradient boosting [XGBoost] (T. Chen et al. 2016), and Bayesian additive regression trees [BART] (Chipman et al. 2010). The linear prediction algorithms were generalized additive models (Hastie et al. 1990) using thin plate splines (Wood 2003), lasso (Tibshirani 1996), logistic regression, and ordinary least squares (OLS).

### 2.7 Benchmarks

The performance of complex algorithms should be contextualized by thoroughly comparing to simpler alternative approaches or benchmarks. If the benchmark algorithms can achieve similar performance then the statistical machine learning algorithms are unnecessary. The improvement of a novel prediction method over standard benchmarks is known as the *skill* of the prediction method (Brier 1950; F. Sanders 1963) or incremental value. In clinical prediction the primary alternatives to ML are less flexible estimators, including logistic regression, OLS, individual decision trees, and stratification on key clinical predictors. We tested each of these options, where key predictors were peak troponin, qualitative ECG reading, EDACS score, and HEART score. As a complement to stratification, we also evaluated logistic regression and decision trees when restricted to these key predictors.

### 2.8 SuperLearner stacked ensembling

When comparing multiple algorithms an initial choice is to use cross-validation to select the algorithm with the best out-of-sample performance. This honest evaluation procedure allows comparing simpler linear models to complex (e.g. tree-based) ones, such that the appropriate amount of flexibility is chosen for the given dataset. A more nuanced decision would be to consider a weighted average of multiple algorithms - creating a team of algorithms whose contribution to the prediction is based on optimizing out-of-sample performance. That is the nature of stacked ensembles (Breiman 1996; Wolpert 1992), sometimes referred to as the Super Learner algorithm (van der Laan, Polley, et al. 2007). Rather than restrict our prediction machine to a single algorithm, we created a weighted average across all tested algorithms, and estimated weights based on an optimization goal so that they minimize a chosen performance statistic on test data. This ensembling procedure performs asymptotically as well or better than the individual estimators (ibid.). We chose to optimize the mean-squared error [MSE] (i.e. Brier score) in our ensemble, using convex weights based on a non-negative least squares meta-learner. Optimizing on MSE incorporates both discrimination and calibration goals for the ensemble (Murphy et al. 1977). A convex combination of algorithm weights ensures that predictions fall within the convex hull of the constituent learners, while inducing sparsity - i.e. algorithms can have zero weight.

### 2.9 Hyperparameter tuning

Hyperparameter settings adjust an algorithm’s characteristics to a dataset’s sample size, number of predictor variables, measurement error, sparsity in predictor relevance, and correlation structure. The performance gain from hyperparameter tuning is referred to as the *tunability* of an algorithm (Probst, Boulesteix, et al. 2019). RF, for example, works well with default hyperparameters but can benefit from hyperparameter tuning to constrain overfitting (Probst, Wright, et al. 2019; Segal et al. 2011).

We adopted *nested ensembling* as our hyperparameter tuning strategy for RF, XGBoost, and individual decision trees (see eMethods). Much as using a weighted average of different algorithms may be preferable to selecting the single best-performing algorithm, a weighted ensemble of hyperparameter settings for a given algorithm may yield improved performance compared to selecting a single hyperparameter configuration. We created small grids of hyperparameter configurations and estimated a SuperLearner ensemble for a given algorithm in which the ensemble weights selected the hyperparameter settings that maximized out-of-sample performance. This ensemble of hyperparameter settings could potentially select a single configuration due to the sparsity induced by the convex combination, or the optimization could distribute the weighting across multiple configurations if such a weighting improved performance over a single selected configuration. The nesting constrains the learners that are analyzed in the outer SuperLearner ensemble, increasing meta-learning stability (i.e. allocation of weights in the convex combination) and easing interpretability.

### 2.10 Evaluation

We evaluated risk prediction models based on their discrimination, calibration, and clinical utility. Nested cross-validation with 5 folds was used to conduct the discrimination and non-visual calibration analyses. The net benefit and calibration visualization analyses were conducted by stacking the five external test sets. While bootstrap estimation has been promoted for evaluation of clinical prediction models (Austin and Tu 2004; Steyerberg et al. 2001), recent work has shown that the bootstrap can be biased for evaluating the performance of highly adaptive ML algorithms estimators such as random forests (Benkeser et al. 2019).

#### 2.10.1 Discrimination

We chose area under the precision-recall curve (PR-AUC, also known as average precision [AP]) as our primary performance metric for evaluating discrimination, because it highlights performance differences that may be obscured by ROC-AUC with imbalanced data (Cook 2007; Saito et al. 2015). The PR-AUC compares positive predictive value to sensitivity across thresholds, focusing on model performance for the rare positive MACE cases (Davis et al. 2006). We included area under the receiver operating characteristic curve (ROC-AUC or the c-statistic) as our secondary performance metric, which remains popular and interpretable (Janssens et al. 2020). The downside of ROC-AUC is that its reliance on the false positive rate gives the model “credit” for predicting negative cases correctly (true negatives), which is not challenging due to the rarity of MACE (Adhikari et al. 2021). As an exploratory metric we also estimated the adjusted Brier score (index of prediction accuracy) which integrates discrimination and calibration into a single metric (Kattan et al. 2018). We did not conduct a reclassification analysis due to recognized limitations (Hilden et al. 2014; Kerr et al. 2014; Leening et al. 2014; Pepe et al. 2015).

#### 2.10.2 Calibration

Our clinical use case was centered on a risk threshold of 0.5% to classify patients as “low risk” in order to qualify for early discharge. With that clinical goal it was essential to compare the model’s predicted risks to the observed risks, i.e. its *calibration* (Lichtenstein et al. 1981) - also known as reliability (Brier 1950; Murphy et al. 1977) or external correspondence (Yates 1982). We assessed the calibration of predicted probabilities in three ways: 1) calibration curve visualization with lowess-smoothing and decile groups, 2) calculating the average absolute value between the predicted risk and the lowess-smoothed observed risk, known as the mean absolute prediction error [MAPE] (van Smeden et al. 2019) or integrated calibration index [ICI] (Austin and Steyerberg 2019), and 3) calculation of the index of prediction accuracy (IPA), a transformation of the Brier score (Kattan et al. 2018). We did not conduct a Hosmer-Lemeshow group-based calibration test due to its recognized limitations and recommendations against its use (Kramer et al. 2007; Van Calster et al. 2019).

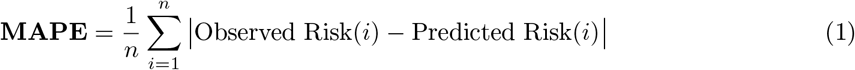

#### 2.10.3 Clinical utility

Although discrimination and calibration are fundamental characteristics of model performance, they do not directly assess the impact of the model on patient care when used for clinical decision support (Vickers, Van Calster, et al. 2016). The planned clinical use of the prediction model was first to assess eligibility for early discharge among low-risk patients (< 0.5%). Accurately estimating the risk of MACE for patients would allow those low-risk patients to be discharged and avoid additional unnecessary workup (overtreatment) and would free up resources (clinical attention, testing capacity, etc.) for higher risk patients.

Our model needed to balance two trade-offs: 1) false negatives where a patient was identified as low-risk but did later have a MACE event, and 2) false positives in which patients were believed to be above the given threshold but who did not later have a MACE event. Errors in the first category have a greater cost than those in the second category, because there is a greater potential detriment to those patients who were discharged early but whose true risk exceeded the threshold. Patients incorrectly estimated to be above the risk threshold, but who are truly low risk, have comparatively lesser costs of additional workup, use of clinical resources, and potential for overtreatment.

This balancing of trade-offs based on a clinical decision threshold can be evaluated using net benefit analysis of decision curves (Vickers and Elkin 2006; Vickers, Van Calster, et al. 2016). The net benefit of a model or strategy at a given threshold is (Vickers and Elkin 2006):

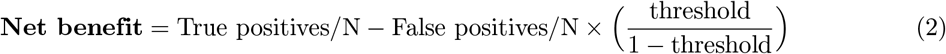

The net benefit formula reflects a relative weighting of true positives versus false positives, which is the threshold / (1 - threshold) term that scales the percentage of false positives. As has been described previously (ibid.), a particular decision threshold inherently embeds a relative weighting. In our case a threshold of 0.5% corresponds to a weight of 0.5% / 99.5% = 0.00503; if we take the inverse that means that a false negative (missed MACE) is 199 times as bad as a false positive (overtreatment). In contrast, a threshold of 2% implies that a missed MACE is only 49 times as bad as further workup for a patient who will not go on to have a MACE.

In a “rule out” scenario where a threshold is applied to a patient’s estimated risk to determine eligibility for early discharge, one can compare benefit-adjusted *net reduction in unnecessary interventions* [NRUI] curves across clinically relevant decision thresholds, scaled to 100 patients for ease of use (ibid.):

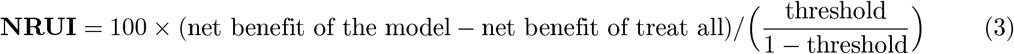

This form of net benefit analysis answers the question: relative to a naive strategy of treating all patients, how many patients can the risk model remove from treatment at a given cutoff threshold, after adjusting for the harm caused by some of those removed patients going on to have a MACE event (false negatives)?

### 2.11 Interpretability

Beyond the statistical performance of a clinical prediction, it is helpful to explain how a model generates its predictions. Interpretability is desirable first because it can provide evidence that the model is working as expected, which can improve the trustworthiness of its predictions for clinicians, patients, or collaborators. Interpretation may also lead to scientific insights about how predictors are related to the outcome, which could be conceptualized as causal pathways, data generating processes, or biological mechanisms. Interpretation can further inform the data export and cleaning processes, such as identifying extreme values, data entry errors, or outliers, or suggesting additional predictor variables to incorporate into the model.

We focus on two complementary forms of model interpretability: *variable predictive importance ranking* and *accumulated local effect plots*, as described below. Prediction-oriented variable importance rankings order the predictor variables by their contribution to a model’s prediction, providing evidence as to which predictors were relied upon the most by the algorithm. Such rankings could be used as a form of confirmatory analysis if a hypothesized ranking were created prior to data analysis, which could formally identify predictors that differed from their expected importance.

It may also be helpful to understand how a model’s prediction varies over the values of individual predictors, particularly continuous predictors with a wide range or large number of unique values. Partial dependence plots (PDPs) as proposed by Friedman (2001) provide this type of interpretability, but they can yield flawed results because they make a key unrealistic assumption that features are statistically independent of each other (Molnar 2020, p. 5.1.3). Accumulated local effect (ALE) plots avoid that limitation of PDPs by counterfactually modifying observations that lie within a nearby kernel neighborhood of the current predictor’s value of interest (Apley et al. 2019). We visualize the contribution of high-importance continuous variables using ALE plots.

## 3 Results

### 3.1 Model performance

#### 3.1.1 Discrimination

Figure 1a displays the precision-recall area under the curve (PR-AUC) and 95% confidence intervals for each combination of features and estimation algorithm. The MACE mean on the training sample represents the baseline PR-AUC (1.88%). The SuperLearner ensemble achieved the highest estimated PR-AUC (0.151, 95% CI [0.141, 0.160]), followed by RF with hyperparameter tuning (0.149, [0.136, 0.162]), the default RF (0.147, [0.135, 0.160]), and the tuned XGBoost (0.139, [0.134, 0.144]). By comparison the PR-AUC for logistic regression was 0.122 [0.114, 0.129], noticeably lower than the ensemble. Point estimates and confidence intervals are listed in eTable 2.

**Figure 1.**
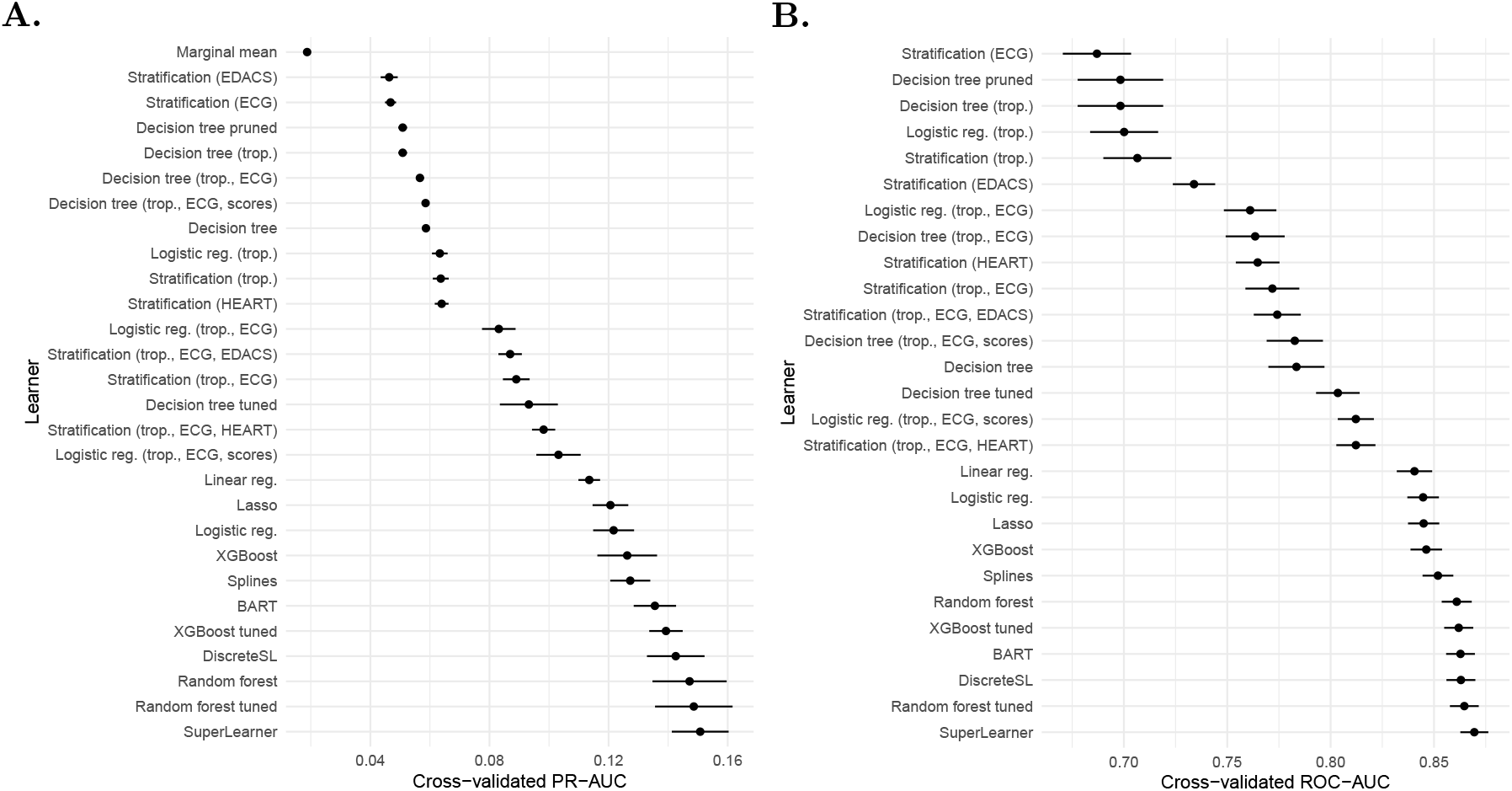
Comparison of cross-validated discriminative performance using a) PR-AUC metric and b) ROC-AUC, with 95% confidence intervals.

Cross-validated ROC-AUC is displayed in Figure 1b (LeDell et al. 2015). The SuperLearner ensemble again achieved the highest performance (ROC-AUC = 0.870, 95% CI [0.863, 0.876]), followed by tuned RF (0.865, [0.858, 0.872]), BART (0.863, [0.856, 0.870]), and tuned XGBoost (0.862, [0.854, 0.869]). The ROC-AUC for logistic regression (0.845, [0.837, 0.852]) was significantly lower than the ensemble. Point estimates and confidence intervals are listed in eTable 3.

We reviewed the distribution of learner weights in the SuperLearner ensemble to assess which algorithms were most useful (Table 1). Four algorithms were always incorporated into the ensemble: default RF (average weight = 0.36), BART (average weight = 0.20), default XGBoost (average weight = 0.20), and tuned RF (average weight = 0.11). The remaining 4 learners were sometimes incorporated into the ensemble, with average weights ranging from 0.04 to 0.007 and a maximum individual weight of 0.07.

**Table 1.** Distribution of algorithm weights across ensemble cross-validation replications

| # | Learner | Mean | SD | Min | Max |
| --- | --- | --- | --- | --- | --- |
| 1 | Random forest | 0.3620 | 0.0847 | 0.2393 | 0.4401 |
| 2 | BART | 0.2046 | 0.0425 | 0.1291 | 0.2288 |
| 3 | XGBoost | 0.1986 | 0.0312 | 0.1481 | 0.2298 |
| 4 | Random forest tuned | 0.1146 | 0.1153 | 0.0019 | 0.2792 |
| 5 | Logistic reg. | 0.0449 | 0.0268 | 0.0000 | 0.0692 |
| 6 | Splines | 0.0408 | 0.0364 | 0.0000 | 0.0790 |
| 7 | XGBoost tuned | 0.0276 | 0.0618 | 0.0000 | 0.1382 |
| 8 | Logistic reg. (trop., ECG, scores) | 0.0069 | 0.0103 | 0.0000 | 0.0239 |

#### 3.1.2 Calibration

We visually compared the predicted risk of the SuperLearner ensemble to the lowess-smoothed observed risk in Figure 2. The median predicted risk was 0.62%, with a first quartile of 0.2% and third quartile of 2%. Given those low risk levels, it would be best to “zoom in” our visual calibration review to that region. We show a zoomed calibration plot as Figure 2a. We further include a exponential-scale calibration plot (Figure 2b) with calibration confidence intervals after grouping patients into 10 groups based on predicted risk, consistent with TRIPOD guideline recommendations (Collins et al. 2015). Due to the low rate of MACE the exponential scaling of axes allows easier comparison across the probability range, although it is less visually intuitive due to the shifting of scales.

**Figure 2.**
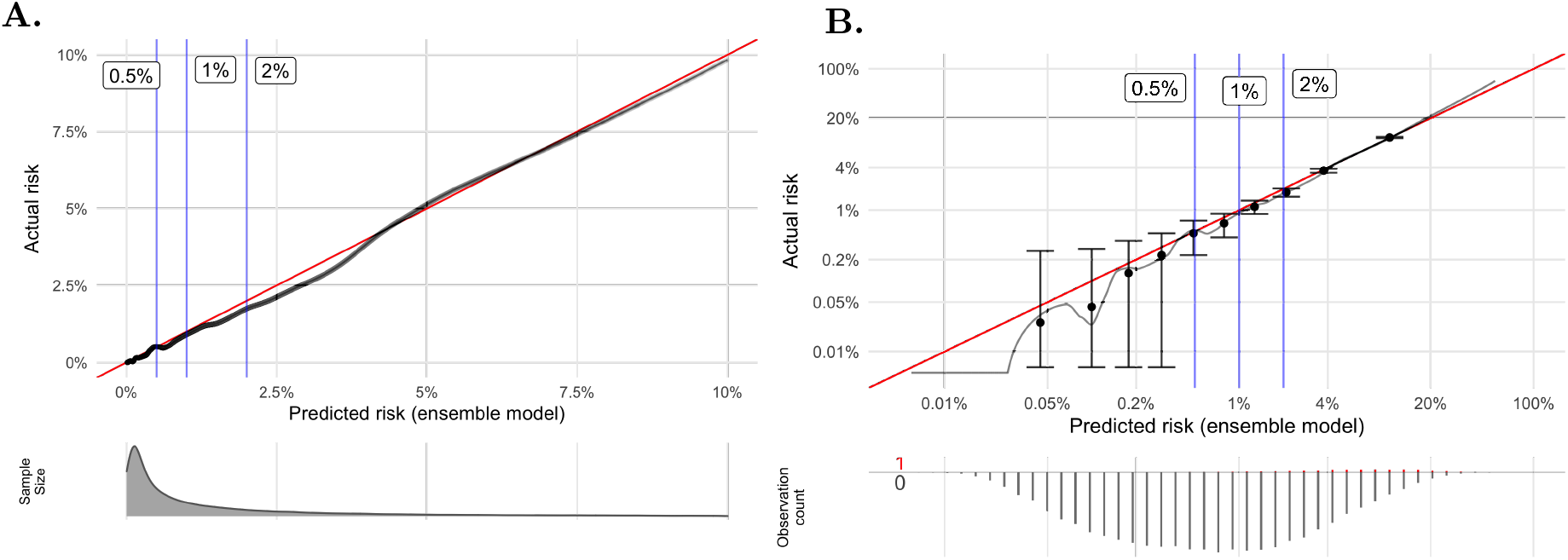
Calibration plot comparing predicted risk to observed risk for the ensemble model. Clinical thresholds of 0.5%, 1%, or 2% risk are noted by blue vertical lines. The red line is the target calibration in which predicted risk is equal to observed risk. A) linear scale, b) log scale.

We found a mean absolute prediction error (MAPE) of 0.20% with a lowess smoothing span of 0.05 (low smoothing), and an MAPE of 0.15% with a smoothing span of 0.20 (moderate smoothing), for a typical miscalibration of about 0.17 percentage points. The supplemental information includes results for the index of prediction accuracy, Brier score, and decile-grouped calibration (eTables 5-7).

#### 3.1.3 Clinical utility

The ensemble model dominated the net reduction in interventions curves across all clinically relevant decision thresholds between 0% and 4% (Figure 3). This finding demonstrates that the ensemble model would lead to the best patient care among the evaluated options, and is therefore an ethical imperative to bring into clinical practice. The relative improvement for the ensemble model, as well as RF and logistic regression, was particularly high at conservative decision thresholds between 0% and 0.6%, when compared to the risk scores or a single decision tree. At our primary threshold of 0.5%, the ensemble model avoided 32 net interventions per 100 patients, compared to 29 for random forest, 25 for logistic regression, 14 for HEART, 3 for EDACS, and 2 for a single decision tree (see eTable 8).

**Figure 3.**
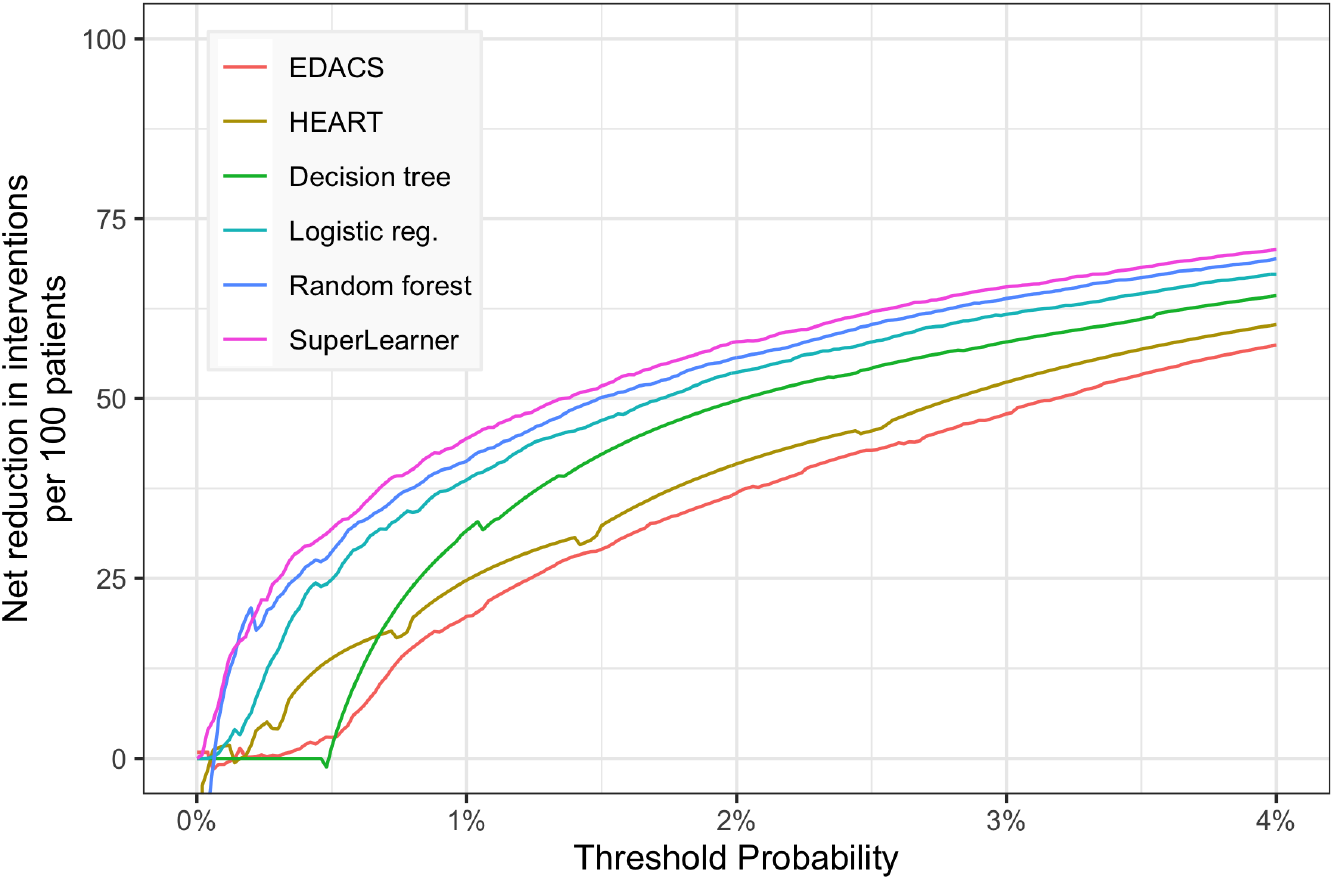
Clinical utility evaluation via benefit-adjusted net reduction in unnecessary interventions per 100 patients.

#### 3.1.4 Missing data imputation

Compared to median/mode imputation, GLRM-based imputation yielded an average reduction in reconstruction error of 90% (eTable 4), suggesting a beneficial ability to capture missingness patterns.

### 3.2 Interpretation

#### 3.2.1 Variable importance ranking

Our final model is complex: it is a weighted average of multiple RF models combined with XGBoost, BART, spline, and logistic regression models, and a stratified estimator. We provide predictive variable importance rankings for the top two estimation algorithms: RF and XGBoost (Table 2).

**Table 2.** Variable importance rankings

| A. Random Forest importance ranking |  | B. XGBoost importance ranking |  |
| --- | --- | --- | --- |
| Variable | Mean<br>Decrease<br>Accuracy (%) | Variable | Gain |
| 1. Age | 0.262 | 1. Peak troponin | 0.3288 |
| 2. EDACS | 0.188 | 2. HEART | 0.1591 |
| 3. HEART | 0.117 | 3. High EDACS | 0.0712 |
| 4. CAD | 0.117 | 4. EDACS | 0.0457 |
| 5. Troponin 3HV | 0.111 | 5. High HEART | 0.0444 |
| 6. LDL | 0.104 | 6. ECG | 0.0415 |
| 7. Total Cholesterol | 0.102 | 7. Peak pulse | 0.0320 |
| 8. Missing Triglycerides | 0.083 | 8. Age | 0.0282 |
| 9. Missing Total Cholesterol | 0.080 | 9. BMI | 0.0234 |
| 10. Missing LDL | 0.076 | 10. SBP | 0.0217 |
| 11. Missing HDL | 0.073 | 11. Myocardial infarction | 0.0210 |
| 12. Pulse | 0.071 | 12. CAD | 0.0198 |
| 13. Peak pulse | 0.070 | 13. Aortic athero. | 0.0175 |
| 14. BMI | 0.051 | 14. Troponin 3HV | 0.0165 |
| 15. Diabetes | 0.047 | 15. HDL | 0.0159 |
| 16. Hypertension | 0.045 | 16. Respiration | 0.0135 |
| 17. HDL | 0.044 | 17. O2 saturation | 0.0131 |
| 18. HbA1c | 0.043 | 18. GFR | 0.0130 |
| 19. Peak troponin | 0.041 | 19. Exertion | 0.0127 |
| 20. Triglycerides | 0.039 | 20. Lowest SBP | 0.0091 |

The rankings differ between the two algorithms, supporting the hypothesis that an ensemble of multiple algorithms could achieve better performance than selecting a single estimation algorithm. Both algorithms place high emphasis on the EDACS and HEART risk scores (as well as binarized versions for XGBoost), demonstrating the benefit of including those scores along with the underlying predictors. Different versions of the cardiac troponin predictor are emphasized by the two algorithms: random forest focuses on 3-hour troponin whereas XGBoost focuses on peak troponin. ECG reading is emphasized by XGBoost but not random forest. Age remains an important predictor, particularly for random forest, despite being a component of the risk scores - highlighting its status as a fundamental predictor of cardiac risk. Both algorithms make use of lipid profile predictors, BMI, and vital signs which are not included in the existing risk scores. The random forest incorporates missingness indicators from lipid panel predictors. Pain-related characteristics sourced from clinical notes are notably not top predictors, a difference from prior MACE prediction work (Amsterdam et al. 2010).

#### 3.2.2 Accumulated local effects

Based on the rankings from the variable importance analyses, we generated accumulated local effect plots (Figure 4) to visualize the conditional relationship of top continuous predictors to the model’s prediction, across their range of values. We can see clustering of the functional relationship for groups of variables (age, EDACS, and HEART; lipid predictors), as well as the general lack of simple linear relationships between the predictors and overall risk.

**Figure 4.**
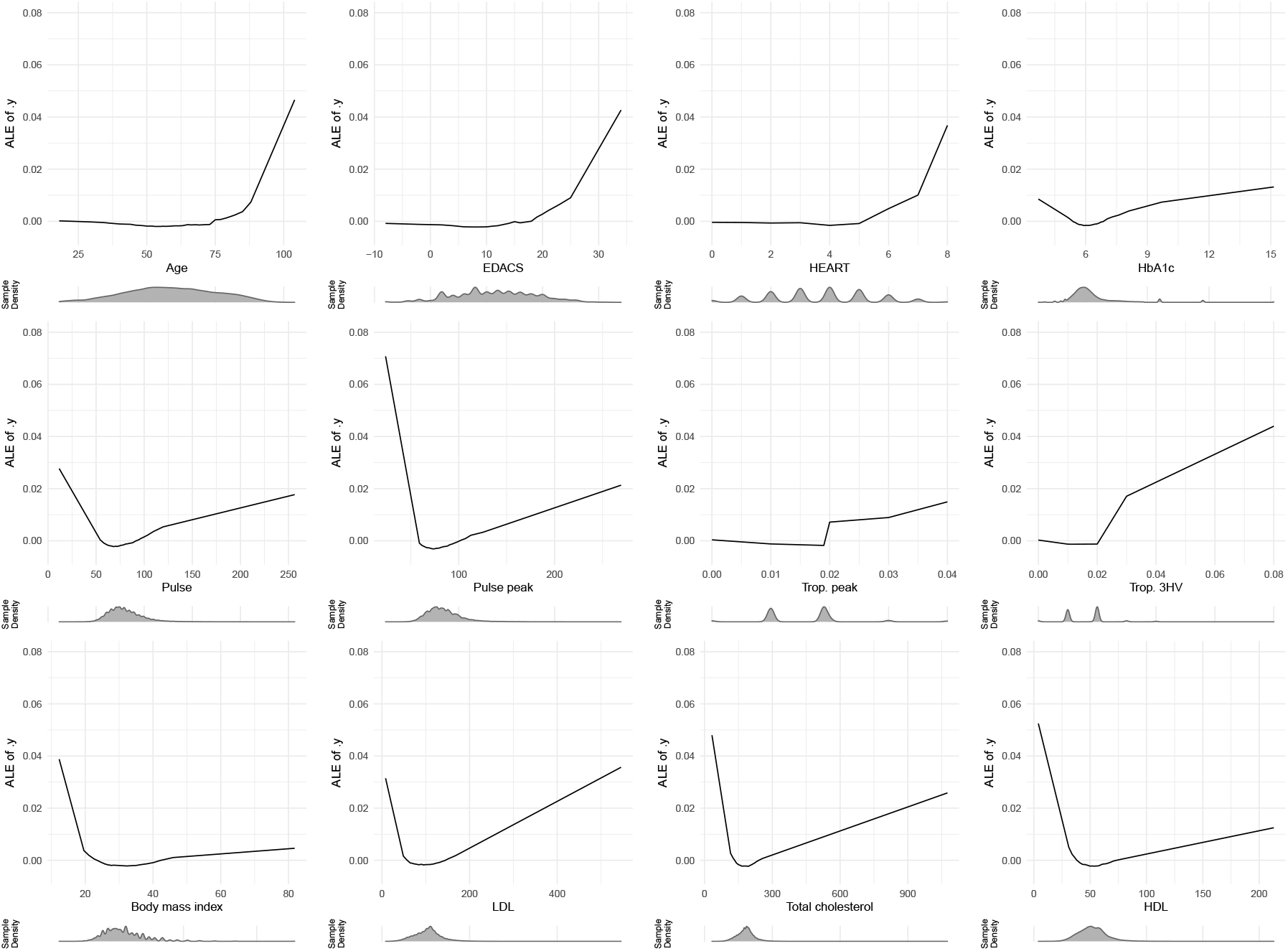
Accumulated local effect plots of key continuous predictors.

## 4 Discussion

It remains debated whether machine learning methods can exhibit statistically and substantively significant benefits for risk prediction compared to logistic regression, decision trees, or additive risk scores (Goldstein, Navar, and Carter 2016; Goldstein, Navar, Pencina, et al. 2016; R. Khera et al. 2021). A recent meta-analysis, for example, did not find systematic benefit from machine learning in comparison to logistic regression (Christodouloua et al. 2019). Yet there is also optimism about the potential for artificial intelligence methods in medicine (He et al. 2019) in general, as well as cardiology (Johnson et al. 2018) and chest pain risk stratification (N. Liu, Chee, et al. 2021; N. Liu, Koh, et al. 2014; Stewart et al. 2021; Zhang et al. 2020) specifically. As long as cross-validation is used as a fair evaluation procedure, the debate is a false dichotomy because we can test and select the algorithm(s) with the appropriate amount of flexibility (bias-variance trade-off) for the signal in the current dataset (van der Laan and Dudoit 2003). Moreover, cross-validated discriminative performance should not be the focal metric for clinical risk modeling: net patient benefit is our ultimate goal, which implicitly focuses on calibration around a clinical decision threshold.

It was important to extract a broader set of granular predictor variables than were used by existing scores, empowering ML methods to capture multi-variable, nonlinear interactions that are missed by linear or additive approaches, perhaps relevant only to certain subgroups of patients. ML may surface novel predictors that have been missed by existing scores or the broader literature, or whose predictive impact was too small, in too complex a form, or underrepresented in terms of sample size to be detected by non-ML methods. The expansion of electronic health records (EHRs) also makes broader covariate collection more feasible and relevant than was possible prior to EHRs, while enabling more granular measurement of variables (E. H. Kennedy et al. 2013).

A complementary strategy was extracting predictors on fine-grained scales, enabling ML to detect novel cut-points or thresholds that improve performance. Variables should be kept as their original continuous measurements rather than dichotomized or discretized into qualitative levels (Senn 2005). For example, dichotomizing BMI into an indicator of high-BMI or the absence of high-BMI loses information compared to the original continuous measurement, and a single threshold may not be optimal for all subgroups or regions of risk. A benefit of ML is that it can identify thresholds in a data-adaptive way, allowing it to better approximate unknown or ill-understood physiological processes.

## 4.1 Future work

The next evaluation step is to externally validate the ensemble model (Moons et al. 2012). This might include cohorts at the current study location (temporal validation) or cohorts sourced from other regions or EHRs (geographical or institutional validation) (ibid.). Cardiac risk assessment will likely continue to shift towards use of high sensitivity troponin (hsTn) assays, which alone perform as well as the HEART and EDACS risk scores. Incorporating such assays into this analytical system could enhance discriminatory power for low risk patients, and improve the granularity of risk assessment for patients with elevated hsTn values. However, because the model uses a wide range of predictor variables in addition to troponin, the incremental value of a higher sensitivity measurement may be limited (M. P. Than et al. 2019).

While technical extensions are possible (detailed in the eMethods), bringing this technology to the bedside is the core remaining task. A real-time clinical decision support tool requires automated extraction and preparation of the predictors for a patient (including possible in-clinic data entry for recently acquired predictors), running the model to generate the prediction, visualizing or otherwise returning the prediction for usage by medical providers, and confirming continued model accuracy (Sendak et al. 2020). Stakeholder engagement, including user testing, training, and feedback, would be necessary to ensure widespread adoption of the tool (Scheinker et al. 2020).

## 5 Conclusion

We explored the benefit of ML algorithms for predicting major adverse cardiac events in patients with chest pain, combining information from cardiac biomarkers (troponin, ECG), additive risk scores, and EHR-sourced patient predictors. The patient cohort was restricted to a lower prevalence subset where nuanced risk prediction could provide clinical value to patients and providers, despite the statistical challenge of a rarer endpoint. The ML algorithms achieved improved discrimination compared to simpler baselines such as logistic regression, decision trees, or stratification on individual predictors. Combining multiple algorithms into an ensemble estimator yielded the best performance. The result was a well-calibrated ensemble model that dominated patient net benefit analysis across risk thresholds, demonstrating the utility for patient care by reducing unnecessary cardiac workups. Finally, we provided interpretation of how the ensemble’s prediction is generated through two methods: ranking the predictors by their contribution to predictive performance, and visualizing the dose-response effect of continuous predictors with accumulated local effect plots.

## Data Availability

The dataset analyzed during the current study is not publicly available due to it containing protected health information (PHI) that could compromise patient privacy. Requests to access the dataset from qualified researchers trained in human subject confidentiality protocols may be sent to the Kaiser Permanente Northern California Institutional Review Board at.

https://github.com/ck37/chest-pain-risk-prediction

https://github.com/ck37/ck37r

## Acknowledgments

We thank Dustin Ballard, MD, Gabriel Escobar, MD, Alan Go, MD, Oleg Sofrygin, PhD, and Jodi McCloskey, MPH for helpful comments, and Adina Rauchwerger, MPH, and Laura Simon, BS for project management.

## Code availability

This study’s analysis code is available at https://github.com/ck37/chest-pain-risk-prediction, with supporting functions in the open source R package ck37r (C. J. Kennedy 2020).

## Author contributions

Study conception and design: Kennedy, Mark, Reed, Huang

Acquisition of data: Reed, Huang, Mark, Kennedy

Analysis and interpretation of data: Kennedy, Mark, Reed, Huang, Hubbard

Drafting of manuscript: Kennedy

Critical revision: Kennedy, Mark, Reed, Hubbard, Huang, van der Laan

## References

Adhikari, Samrachana, Sharon-Lise Normand, Jordan Bloom, David Shahian, and Sherri Rose (2021). “Revisiting performance metrics for prediction with rare outcomes”. In: Statistical Methods in Medical Research 0.0. PMID: 34468239, p. 09622802211038754. doi: 10.1177/09622802211038754. eprint: https://doi.org/10.1177/09622802211038754. URL: https://doi.org/10.1177/09622802211038754.

Agniel, Denis, Isaac S Kohane, and Griffin M Weber (2018). “Biases in electronic health record data due to processes within the healthcare system: retrospective observational study”. In: Bmj 361.

Agor, Joseph, Osman Y Özaltın, Julie S Ivy, Muge Capan, Ryan Arnold, and Santiago Romero (2019). “The value of missing information in severity of illness score development”. In: Journal of biomedical informatics 97, p. 103255.

Amsterdam, Ezra A, J Douglas Kirk, David A Bluemke, Deborah Diercks, Michael E Farkouh, J Lee Garvey, Michael C Kontos, James McCord, Todd D Miller, Anthony Morise, et al. (2010). “Testing of low-risk patients presenting to the emergency department with chest pain: a scientific statement from the American Heart Association”. In: Circulation 122.17, pp. 1756–1776.

Apley, Daniel W and Jingyu Zhu (2019). “Visualizing the effects of predictor variables in black box supervised learning models”. In: arXiv preprint arXiv:1612.08468.

Austin, Peter C and Ewout W Steyerberg (2019). “The Integrated Calibration Index (ICI) and related metrics for quantifying the calibration of logistic regression models”. In: Statistics in medicine 38.21, pp. 4051–4065.

Austin, Peter C and Jack V Tu (2004). “Bootstrap methods for developing predictive models”. In: The American Statistician 58.2, pp. 131–137.

Benkeser, David, Maya Petersen, and Mark J van der Laan (2019). “Improved small-sample estimation of nonlinear cross-validated prediction metrics”. In: Journal of the American Statistical Association, pp. 1–16.

Bonaca, Marc, Benjamin Scirica, Marc Sabatine, Anthony Dalby, Jindrich Spinar, Sabina A Murphy, Peter Jarolim, Eugene Braunwald, and David A Morrow (2010). “Prospective evaluation of the prognostic implications of improved assay performance with a sensitive assay for cardiac troponin I”. In: Journal of the American College of Cardiology 55.19, pp. 2118–2124.

Breiman, Leo (1996). “Stacked regressions”. In: Machine learning 24.1, pp. 49–64.

Breiman, Leo (2001). “Random forests”. In: Machine learning 45.1, pp. 5–32.

Brier, Glenn W (1950). “Verification of forecasts expressed in terms of probability”. In: Monthly weather review 78.1, pp. 1–3.

Chen, Tianqi and Carlos Guestrin (2016). “Xgboost: A scalable tree boosting system”. In: Proceedings of the 22nd acm sigkdd international conference on knowledge discovery and data mining, pp. 785–794.

Chipman, Hugh A, Edward I George, Robert E McCulloch, et al. (2010). “BART: Bayesian additive regression trees”. In: The Annals of Applied Statistics 4.1, pp. 266–298.

Christodouloua, Evangelia, MA Jie, Gary S Collins, Ewout W Steyerberg, Jan Y Verbakel, Ben van Calster, et al. (2019). “A systematic review shows no performance benefit of machine learning over logistic regression for clinical prediction models”. In: Journal of clinical epidemiology.

Collins, Gary S, Johannes B Reitsma, Douglas G Altman, and Karel GM Moons (2015). “Transparent reporting of a multivariable prediction model for individual prognosis or diagnosis (TRIPOD): the TRIPOD statement”. In: British Journal of Surgery 102.3, pp. 148–158.

Cook, Nancy R (2007). “Use and misuse of the receiver operating characteristic curve in risk prediction”. In: Circulation 115.7, pp. 928–935.

Davis, Jesse and Mark Goadrich (2006). “The relationship between Precision-Recall and ROC curves”. In: Proceedings of the 23rd international conference on Machine learning, pp. 233–240.

De Lemos, James A, Mark H Drazner, Torbjorn Omland, Colby R Ayers, Amit Khera, Anand Rohatgi, Ibrahim Hashim, Jarett D Berry, Sandeep R Das, David A Morrow, et al. (2010). “Association of troponin T detected with a highly sensitive assay and cardiac structure and mortality risk in the general population”. In: Jama 304.22, pp. 2503–2512.

Friedman, Jerome H (2001). “Greedy function approximation: a gradient boosting machine”. In: Annals of statistics, pp. 1189–1232.

Goldstein, Benjamin A, Ann Marie Navar, and Rickey E. Carter (2016). “Moving beyond regression techniques in cardiovascular risk prediction: applying machine learning to address analytic challenges”. In: European Heart Journal 38.23, pp. 1805–1814. ISSN: 0195-668X. doi: 10.1093/eurheartj/ehw302.

Goldstein, Benjamin A, Ann Marie Navar, Michael J Pencina, and John P A Ioannidis (2016). “Opportunities and challenges in developing risk prediction models with electronic health records data: a systematic review”. In: Journal of the American Medical Informatics Association 24.1, pp. 198–208. ISSN: 1067-5027. doi: 10.1093/jamia/ocw042.

Greenslade, Jaimi H, Edward W Carlton, Christopher Van Hise, Elizabeth Cho, Tracey Hawkins, William A Parsonage, Jillian Tate, Jacobus Ungerer, and Louise Cullen (2018). “Diagnostic accuracy of a new high-sensitivity troponin I assay and five accelerated diagnostic pathways for ruling out acute myocardial infarction and acute coronary syndrome”. In: Annals of emergency medicine 71.4, pp. 439–451.

Groenwold, Rolf HH (2020). “Informative missingness in electronic health record systems: the curse of knowing”. In: Diagnostic and prognostic research 4.1, pp. 1–6.

Hastie, Trevor J and Robert J Tibshirani (1990). Generalized additive models. Vol. 43. CRC press.

He, Jianxing, Sally L Baxter, Jie Xu, Jiming Xu, Xingtao Zhou, and Kang Zhang (2019). “The practical implementation of artificial intelligence technologies in medicine”. In: Nature medicine 25.1, p. 30.

Hilden, Jørgen and Thomas A Gerds (2014). “A note on the evaluation of novel biomarkers: do not rely on integrated discrimination improvement and net reclassification index”. In: Statistics in medicine 33.19, pp. 3405–3414.

Janssens, A Cecile J W and Forike K Martens (2020). “Reflection on modern methods: Revisiting the area under the ROC Curve”. In: International Journal of Epidemiology. dyz274. ISSN: 0300-5771. doi: 10.1093/ije/dyz274.

Johnson, Kipp W, Jessica Torres Soto, Benjamin S Glicksberg, Khader Shameer, Riccardo Miotto, Mohsin Ali, Euan Ashley, and Joel T Dudley (2018). “Artificial intelligence in cardiology”. In: Journal of the American College of Cardiology 71.23, pp. 2668–2679.

Kattan, Michael W and Thomas A Gerds (2018). “The index of prediction accuracy: an intuitive measure useful for evaluating risk prediction models”. In: Diagnostic and prognostic research 2.1, p. 7.

Kennedy, Chris J (2020). ck37r: Chris Kennedy’s R toolkit. URL: https://github.com/ck37/ck37r.

Kennedy, Edward H, Wyndy L Wiitala, Rodney A Hayward, and Jeremy B Sussman (2013). “Improved cardiovascular risk prediction using nonparametric regression and electronic health record data”. In: Medical care 51.3, p. 251.

Kerr, Kathleen F, Zheyu Wang, Holly Janes, Robyn L McClelland, Bruce M Psaty, and Margaret S Pepe (2014). “Net reclassification indices for evaluating risk-prediction instruments: a critical review”. In: Epidemiology (Cambridge, Mass.) 25.1, p. 114.

Khera, Rohan, Julian Haimovich, Nathan C Hurley, Robert McNamara, John A Spertus, Nihar Desai, John S Rumsfeld, Frederick A Masoudi, Chenxi Huang, Sharon-Lise Normand, et al. (2021). “Use of machine learning models to predict death after acute myocardial infarction”. In: JAMA cardiology.

Kontos, Michael C, Deborah B Diercks, and J Douglas Kirk (2010). “Emergency department and office-based evaluation of patients with chest pain”. In: Mayo Clinic Proceedings. Vol. 85. 3. Elsevier, pp. 284–299.

Kramer, Andrew A and Jack E Zimmerman (2007). “Assessing the calibration of mortality benchmarks in critical care: The Hosmer-Lemeshow test revisited”. In: Critical care medicine 35.9, pp. 2052–2056.

LeDell, Erin, Maya Petersen, and Mark van der Laan (2015). “Computationally efficient confidence intervals for cross-validated area under the ROC curve estimates”. In: Electronic journal of statistics 9.1, p. 1583.

Leening, Maarten JG, Moniek M Vedder, Jacqueline CM Witteman, Michael J Pencina, and Ewout W Steyerberg (2014). “Net reclassification improvement: computation, interpretation, and controversies: a literature review and clinician’s guide”. In: Annals of internal medicine 160.2, pp. 122–131.

Lichtenstein, Sarah, Baruch Fischhoff, and Lawrence D Phillips (1981). Calibration of probabilities: The state of the art to 1980. Tech. rep. Decision Research. Eugene, OR.

Liu, Nan, Marcel Lucas Chee, Zhi Xiong Koh, Su Li Leow, Andrew Fu Wah Ho, Dagang Guo, and Marcus Eng Hock xOng (2021). “Utilizing machine learning dimensionality reduction for risk stratification of chest pain patients in the emergency department”. In: BMC medical research methodology 21.1, pp. 1–13.

Liu, Nan, Zhi Xiong Koh, Junyang Goh, Zhiping Lin, Benjamin Haaland, Boon Ping Ting, and Marcus Eng Hock Ong (2014). “Prediction of adverse cardiac events in emergency department patients with chest pain using machine learning for variable selection”. In: BMC medical informatics and decision making 14.1, pp. 1–9.

Mark, Dustin G, Jie Huang, Uli Chettipally, Mamata V Kene, Megan L Anderson, Erik P Hess, Dustin W Ballard, David R Vinson, and Mary E Reed (2018). “Performance of coronary risk scores among patients with chest pain in the emergency department”. In: Journal of the American College of Cardiology 71.6, pp. 606–616.

Mark, Dustin G, Jie Huang, Mamata V Kene, Dana R Sax, Dale M Cotton, James S Lin, Sean C Bouvet, Uli K Chettipally, Megan L Anderson, Ian D McLachlan, et al. (2021). “Prospective Validation and Comparative Analysis of Coronary Risk Stratification Strategies Among Emergency Department Patients With Chest Pain”. In: Journal of the American Heart Association 10.7, e020082.

Molnar, Christoph (2020). Interpretable Machine Learning. Lulu. com.

Moons, Karel GM, Andre Pascal Kengne, Diederick E Grobbee, Patrick Royston, Yvonne Vergouwe, Douglas G Altman, and Mark Woodward (2012). “Risk prediction models: II. External validation, model updating, and impact assessment”. In: Heart 98.9, pp. 691–698.

Murphy, Allan H and Robert L Winkler (1977). “Reliability of subjective probability forecasts of precipitation and temperature”. In: Journal of the Royal Statistical Society: Series C (Applied Statistics) 26.1, pp. 41–47.

Pepe, Margaret S, Jing Fan, Ziding Feng, Thomas Gerds, and Jorgen Hilden (2015). “The net reclassification index (NRI): a misleading measure of prediction improvement even with independent test data sets”. In: Statistics in biosciences 7.2, pp. 282–295.

Probst, Philipp, Anne-Laure Boulesteix, and Bernd Bischl (2019). “Tunability: Importance of Hyperparameters of Machine Learning Algorithms.” In: Journal of Machine Learning Research 20.53, pp. 1–32.

Probst, Philipp, Marvin N Wright, and Anne-Laure Boulesteix (2019). “Hyperparameters and tuning strategies for random forest”. In: Wiley Interdisciplinary Reviews: Data Mining and Knowledge Discovery 9.3, e1301.

Rozenholc, Yves, Thoralf Mildenberger, and Ursula Gather (2010). “Combining regular and irregular histograms by penalized likelihood”. In: Computational Statistics & Data Analysis 54.12, pp. 3313–3323.

Rui, P, K Kang, and JJ. Ashman (2016). National Hospital Ambulatory Medical Care Survey: 2016 emergency department summary tables. URL: https://www.cdc.gov/nchs/data/ahcd/nhamcs_emergency/2016_ed_web_tables.pdf.

Sabbatini, Amber K, Brahmajee K Nallamothu, and Keith E Kocher (2014). “Reducing variation in hospital admissions from the emergency department for low-mortality conditions may produce savings”. In: Health affairs 33.9, pp. 1655–1663.

Saito, Takaya and Marc Rehmsmeier (2015). “The precision-recall plot is more informative than the ROC plot when evaluating binary classifiers on imbalanced datasets”. In: PloS one 10.3.

Sanders, Frederick (1963). “On subjective probability forecasting”. In: Journal of Applied Meteorology 2.2, pp. 191–201.

Scheinker, David and Margaret L Brandeau (2020). “Implementing analytics projects in a hospital: successes, failures, and opportunities”. In: INFORMS Journal on Applied Analytics 50.3, pp. 176–189.

Schuler, Alejandro, Vincent Liu, Joe Wan, Alison Callahan, Madeleine Udell, David E Stark, and Nigam H Shah (2016). “Discovering patient phenotypes using generalized low rank models”. In: Biocomputing 2016: Proceedings of the Pacific Symposium. World Scientific, pp. 144–155.

Scirica, Benjamin M (2010). “Acute coronary syndrome: emerging tools for diagnosis and risk assessment”. In: Journal of the American College of Cardiology 55.14, pp. 1403–1415.

Segal, Mark and Yuanyuan Xiao (2011). “Multivariate random forests”. In: Wiley Interdisciplinary Reviews: Data Mining and Knowledge Discovery 1.1, pp. 80–87.

Sendak, Mark P, William Ratliff, Dina Sarro, Elizabeth Alderton, Joseph Futoma, Michael Gao, Marshall Nichols, Mike Revoir, Faraz Yashar, Corinne Miller, et al. (2020). “Real-world integration of a sepsis deep learning technology into routine clinical care: implementation study”. In: JMIR medical informatics 8.7, e15182.

Senn, Stephen (2005). “Dichotomania: an obsessive compulsive disorder that is badly affecting the quality of analysis of pharmaceutical trials”. In: Proceedings of the International Statistical Institute, 55th Session, Sydney.

Sisk, Rose, Lijing Lin, Matthew Sperrin, Jessica K Barrett, Brian Tom, Karla Diaz-Ordaz, Niels Peek, and Glen P Martin (2021). “Informative presence and observation in routine health data: A review of methodology for clinical risk prediction”. In: Journal of the American Medical Informatics Association 28.1, pp. 155–166.

Smith, Jennifer N, Jenna M Negrelli, Megha B Manek, Emily M Hawes, and Anthony J Viera (2015). “Diagnosis and management of acute coronary syndrome: an evidence-based update”. In: The Journal of the American Board of Family Medicine 28.2, pp. 283–293.

Sperrin, Matthew, Glen P Martin, Rose Sisk, and Niels Peek (2020). “Missing data should be handled differently for prediction than for description or causal explanation”. In: Journal of Clinical Epidemiology 125, pp. 183–187.

Stewart, Jonathon, Juan Lu, Adrian Goudie, Mohammed Bennamoun, Peter Sprivulis, Frank Sanfillipo, and Girish Dwivedi (2021). “Applications of machine learning to undifferentiated chest pain in the emergency department: A systematic review”. In: PloS one 16.8, e0252612.

Steyerberg, Ewout W, Frank E Harrell Jr, Gerard JJM Borsboom, MJC Eijkemans, Yvonne Vergouwe, and J Dik F Habbema (2001). “Internal validation of predictive models: efficiency of some procedures for logistic regression analysis”. In: Journal of clinical epidemiology 54.8, pp. 774–781.

Than, Martin P, John W Pickering, Yader Sandoval, Anoop SV Shah, Athanasios Tsanas, Fred S Apple, Stefan Blankenberg, Louise Cullen, Christian Mueller, Johannes T Neumann, et al. (2019). “Machine learning to predict the likelihood of acute myocardial infarction”. In: Circulation 140.11, pp. 899–909.

Than, Martin, Dylan Flaws, Sharon Sanders, Jenny Doust, Paul Glasziou, Jeffery Kline, Sally Aldous, Richard Troughton, Christopher Reid, William A Parsonage, et al. (2014). “Development and validation of the E mergency D epartment A ssessment of C hest pain S core and 2 h accelerated diagnostic protocol”. In: Emergency Medicine Australasia 26.1, pp. 34–44.

Than, Martin, Mel Herbert, Dylan Flaws, Louise Cullen, Erik Hess, Judd E Hollander, Deborah Diercks, Michael W Ardagh, Jeffery A Kline, Zea Munro, et al. (2013). “What is an acceptable risk of major adverse cardiac event in chest pain patients soon after discharge from the Emergency Department?: a clinical survey”. In: International journal of cardiology 166.3, pp. 752–754.

Tibshirani, Robert (1996). “Regression shrinkage and selection via the lasso”. In: Journal of the Royal Statistical Society: Series B (Methodological) 58.1, pp. 267–288.

Udell, Madeleine, Corinne Horn, Reza Zadeh, Stephen Boyd, et al. (2016). “Generalized low rank models”. In: Foundations and Trends® in Machine Learning 9.1, pp. 1–118.

Van Calster, Ben, David J McLernon, Maarten van Smeden, Laure Wynants, and Ewout W Steyerberg (2019). “Calibration: the Achilles heel of predictive analytics”. In: BMC medicine 17.1, pp. 1–7.

van der Laan, Mark J and Sandrine Dudoit (2003). “Unified cross-validation methodology for selection among estimators and a general cross-validated adaptive epsilon-net estimator: Finite sample oracle inequalities and examples”. In:

van der Laan, Mark J, Eric C Polley, and Alan E Hubbard (2007). “Super learner”. In: Statistical applications in genetics and molecular biology 6.1.

van Smeden, Maarten, Karel GM Moons, Joris AH de Groot, Gary S Collins, Douglas G Altman, Marinus JC Eijkemans, and Johannes B Reitsma (2019). “Sample size for binary logistic prediction models: beyond events per variable criteria”. In: Statistical methods in medical research 28.8, pp. 2455–2474.

Vickers, Andrew J and Elena B Elkin (2006). “Decision curve analysis: a novel method for evaluating prediction models”. In: Medical Decision Making 26.6, pp. 565–574.

Vickers, Andrew J, Ben Van Calster, and Ewout W Steyerberg (2016). “Net benefit approaches to the evaluation of prediction models, molecular markers, and diagnostic tests”. In: bmj 352.

Wolpert, David H (1992). “Stacked generalization”. In: Neural networks 5.2, pp. 241–259.

Wood, Simon N (2003). “Thin plate regression splines”. In: Journal of the Royal Statistical Society: Series B (Statistical Methodology) 65.1, pp. 95–114.

Yates, J Frank (1982). “External correspondence: Decompositions of the mean probability score”. In: Organizational Behavior and Human Performance 30.1, pp. 132–156.

Zhang, Pei-I, Chien-Chin Hsu, Yuan Kao, Chia-Jung Chen, Ya-Wei Kuo, Shu-Lien Hsu, Tzu-Lan Liu, Hung-Jung Lin, Jhi-Joung Wang, Chung-Feng Liu, et al. (2020). “Real-time AI prediction for major adverse cardiac events in emergency department patients with chest pain”. In: Scandinavian Journal of Trauma, Resuscitation and Emergency Medicine 28.1, pp. 1–7.

