## Supplementary material for "Development of an ensemble machine learning prognostic model to predict 60-day risk of major adverse cardiac events in adults with chest pain"

### eMethods

#### Linear prediction algorithms

Splines have shown competitive performance with tree-based algorithms in prior clinical prediction work due to their ability to identify non-linear, but smooth patterns (Austin 2007). The lasso algorithm (or its generalization the elastic net) is a helpful test of sparsity in the covariates, and a faster & more nuanced variable selection method than best subset or stepwise selection (Hastie et al. 2017). Better performance for lasso compared to logistic regression would indicate that feature selection could be helpful for other algorithms, while equal performance could indicate that the extraction of predictors from the EHR was overly restrictive and should be broadened (i.e. because we are not yet benefiting from penalization).

#### Nested ensemble hyperparameter tuning

The random forest grid consisted of 9 configurations: minimum node size $\in \{5, 20, 60\}$ $\times$ covariates sampled $\in \{4, 8, 16\}$. The number of covariates sampled (i.e. mtry) was based on the formula: $\text{floor}(\{0.5, 1, 2\} \cdot \sqrt{p})$ where p is the total number of covariates. The xgboost grid consisted of 8 configurations: number of trees $\in \{250, 1000\}$ $\times$ maximum tree depth $\in \{2, 4\}$ $\times$ shrinkage $\in \{0.05, 0.2\}$. The decision tree grid consisted of 12 configurations: complexity parameter $\in \{0, 0.01\}$ $\times$ minimum split $\in$ $\{10, 20, 80\}$ $\times$ maximum tree depth $\in \{10, 30\}$.

#### Variable importance details

For the random forest variable importance ranking analysis, the binarized versions of HEART and EDACS risk scores were excluded due to the potential for a masking effect between those predictors and the non-binarized versions of the scores. This risk of a masking effect of correlated predictors is one limitation of permutation-based single-variable importance ranking algorithms (Molnar 2020, Section 7.5).

#### GLRM imputation details

The GLRM hyperparameter settings were chosen through a grid search in which each model was trained on 75% of the data and evaluated on the remaining 25% for accuracy at reconstructing the original observed data matrix. Missingness indicators were not included in the GLRM imputation analysis. Cells with missing data were then replaced with the reconstructed data matrix from GLRM using the optimal settings. Our final GLRM settings were: 50 components, quadratic regularization on X with weight 4, and L1 regularization on Y with weight 24. Here X refers to the reduced components after GLRM transformation, and Y refers to the complementary matrix that transforms those components back to the original covariate space. Multiple imputation was not necessary because our scientific goal was to characterize predictive performance for the unimputed outcome variable, rather than to estimate statistical parameters for covariates that were imputed, such as linear regression coefficients (Sisk et al. 2021; Sperrin et al. 2020).

#### Future technical improvements

We aim to expand the machine learning in several ways in future work. Cross-validation could be conducted using temporal splitting to take into account the loss in performance due to distribution changes over time (Roberts et al. 2017). Additional machine learning algorithms could be tested for benefit, such as LightGBM, CatBoost, extremely randomized trees, multivariate adaptive regression splines, and deep learning for tabular data. The ensemble weighting (metalearner) could directly optimize net benefit. Incorporating feature ranking and selection may benefit the simpler algorithms by removing unhelpful predictors, such as through ensemble methods (Effrosynidis et al. 2021). Feature engineering might be beneficial as well, such as creation of interaction terms or even incorporation of the principal components from the GLRM imputation. Due to computational limitations we were not able to conduct hyperparameter tuning on the BART learner, which likely would provide some performance benefit. We are optimistic that random search or model-based search (e.g. Hyperband) rather than grid search could provide even stronger tuning of algorithm hyperparameters across a higher number of dimensions (Li et al. 2017). Imputation model training and data standardization could be restricted to the training fold, avoiding even unsupervised use of test set data (Jaeger et al. 2020). Evaluation of the GLRM imputation could be further contextualized through cross-validated comparisons to additional imputation methods, especially principal component analysis, k-nearest neighbors, and deep autoencoders, or even stacked ensemble imputation strategies. The variable importance ranking could be extended through a random forest-style permutation importance analysis of the ensemble model, conditional importance measures (Strobl et al. 2007), and through targeted learning methods such as vimp (Williamson et al. 2017) or varimpact (Hubbard et al. 2018). Calibration might be improved through targeted learning-based adjustment (Brooks et al. 2012) or isotonic regression on validation data. Cross-validated estimation of discrimination performance could be improved through cross-validated targeted maximum likelihood estimation (Benkeser et al. 2019).

### Supplemental Figures

#### eFigure 1. Single decision tree benchmark

A single decision tree is a helpful benchmark approach to risk prediction. It is easy to explain and to visualize, and represents a decision-making process that can be conducted by medical staff. It is limited by its simplicity though: decision trees rarely have competitive predictive performance and the selected variables & cutpoints are known to be sensitive to small changes to the dataset. The figure shows an example tree as estimated on our data. Each leaf node shows 1) predicted risk, and 2) the percentage of patients that fall into that node. Here “troponin” refers to peak troponin.

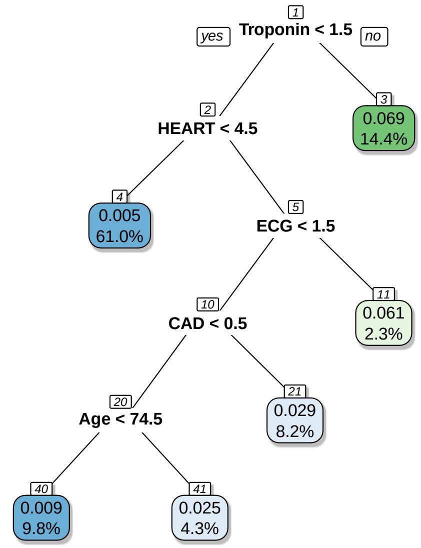


Of particular clinical interest is the blue leaf node (#4) showing a predicted risk of 0.5% and encompassing 61% of the sample. That node represents the lowest-risk group as identified by the decision tree. On first glance the identification of the low-risk group as comprising 61% of patients would seem to improve upon the 50% low-risk result from our complex nested ensemble. However, the 0.5% estimated risk for that leaf node is an average across the large proportion of patients that met the two simple decisions: non-elevated troponin and HEART score $\leq 4$. We know that every such patient doesn't have exactly 0.5% risk - there is a distribution around that average, with some patients have lower risk and others having higher risk. Thus we can see that this low-risk node must suffer from some miscalibration: an unknown proportion of those patients have higher risk than our target 0.5% threshold, and should not be discharged early despite the recommendation of the decision tree.

#### eFigure 2. Random forest convergence plot shows ROC-AUC on out-of-bag data as forest size is increased exponentially

The number of trees is a key hyperparameter for random forest. Breiman (2001) proved that adding additional trees does not lead to overfitting due to the independence of the tree estimators, unlike the sequential fitting of boosting approaches. Yet it remains beneficial to examine the convergence of the random forest's performance to ensure that an adequate number of trees is being used (Probst et al. 2017). To assess performance convergence we analyzed the out-of-bag ROC-AUC for up to 3,000 trees. From visual inspection we can see that only 100 trees would be inadequate, whereas 1,000 - 3,000 trees achieves an ROC-AUC above 0.85.

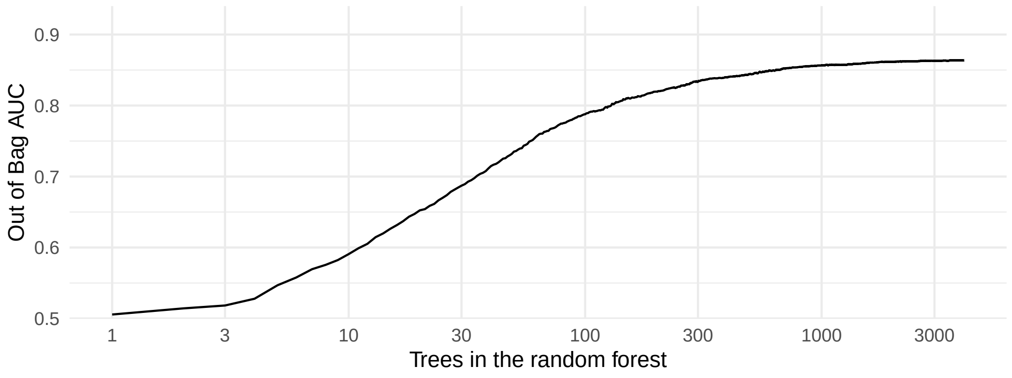


**eFigure 3. Comparison of cross-validated discriminative performance using precision-recall curves for a subset of learners**

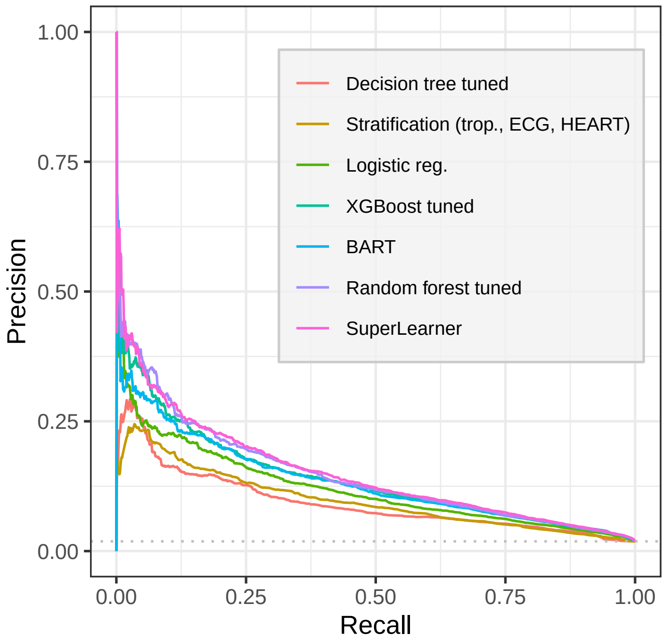

##### eFigure 4. Overall calibration plot comparing predicted risk to observed risk

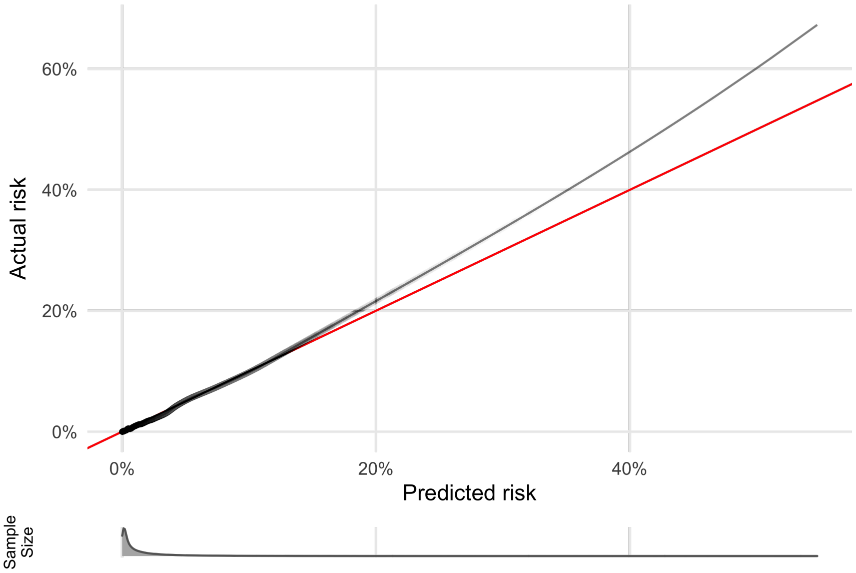


### Supplemental Tables

#### eTable 1. Predictor summary

This table summarizes the predictors used in the model following missing value imputation and histogram condensing of unique values for high-cardinality variables. This table includes 71 total predictors but totals 74 predictors when the race predictor is decomposed from a categorical predictor into 4 indicator predictors.

| | Name | Unique values | Mode | Mean | Median | Minimum | Maximum |
|---|---|---|---|---|---|---|---|
| **Biomarkers** | | | | | | | |
| 1 | ECG | 3 | 0 | 0.44 | 0.00 | 0.00 | 2.00 |
| 2 | Peak troponin | 6 | 0.02 | 0.02 | 0.02 | 0.00 | 0.04 |
| 3 | Troponin 3HV | 29 | 0.02 | 0.02 | 0.02 | 0.00 | 0.08 |
| 4 | Troponin categories | 2 | 1 | 1.14 | 1.00 | 1.00 | 2.00 |
| **Scores** | | | | | | | |
| 5 | EDACS | 41 | 8 | 11.49 | 12.00 | -8.00 | 34.00 |
| 6 | HEART | 9 | 4 | 3.69 | 4.00 | 0.00 | 8.00 |
| 7 | High EDACS | 2 | 0 | 0.34 | 0.00 | 0.00 | 1.00 |
| 8 | High HEART | 2 | 1 | 0.54 | 1.00 | 0.00 | 1.00 |
| **Labs** | | | | | | | |
| 9 | GFR | 96 | 70 | 65.77 | 70.00 | 2.00 | 70.00 |
| 10 | HbA1c | 83 | 6.15 | 6.37 | 6.15 | 3.70 | 15.15 |
| 11 | HDL | 148 | 48 | 51.17 | 50.00 | 4.00 | 213.00 |
| 12 | LDL | 137 | 110.95 | 103.60 | 102.85 | 8.35 | 545.65 |
| 13 | Total Cholesterol | 104 | 186.38 | 180.75 | 180.89 | 32.74 | 1075.26 |
| 14 | Triglycerides | 86 | 4.74 | 4.77 | 4.74 | 3.37 | 8.56 |

**Table S1.** Summary of predictors *(continued)*

| | Name | Unique values | Mode | Mean | Median | Minimum | Maximum |
|---|---|---|---|---|---|---|---|
| **Vitals** | | | | | | | |
| 15 | BMI | 100 | 30.37 | 29.76 | 28.42 | 12.40 | 81.82 |
| 16 | Lowest SBP | 174 | 120 | 121.29 | 120.00 | 42.00 | 244.00 |
| 17 | O2 saturation | 45 | 100 | 98.05 | 98.00 | 16.00 | 100.00 |
| 18 | Obesity | 2 | 0 | 0.42 | 0.00 | 0.00 | 1.00 |
| 19 | Peak pulse | 153 | 77.34 | 84.34 | 81.57 | 23.71 | 269.29 |
| 20 | Pulse | 168 | 74.38 | 80.24 | 78.15 | 11.63 | 256.37 |
| 21 | Respiration | 67 | 18 | 18.10 | 18.00 | 2.00 | 100.00 |
| 22 | SBP | 155 | 145.35 | 144.58 | 142.71 | 55.66 | 269.34 |
| **Demographics** | | | | | | | |
| 23 | Age | 87 | 53 | 59.00 | 59.00 | 18.00 | 104.00 |
| 24 | Male | 2 | 0 | 0.42 | 0.00 | 0.00 | 1.00 |
| 25 | Race | 5 | White | - | - | - | - |
| **Clinical notes** | | | | | | | |
| 26 | Diaphoresis | 3 | 0 | 0.17 | 0.00 | 0.00 | 9.00 |
| 27 | Exertion | 3 | 0 | 0.23 | 0.00 | 0.00 | 9.00 |
| 28 | Pain on inspiration | 3 | 0 | 0.20 | 0.00 | 0.00 | 9.00 |
| 29 | Pain on palpation | 3 | 0 | 0.14 | 0.00 | 0.00 | 9.00 |
| 30 | Radiating pain | 6 | 0 | 0.61 | 0.00 | 0.00 | 9.00 |
| 31 | Sharp pain | 3 | 0 | 0.21 | 0.00 | 0.00 | 9.00 |
| **History** | | | | | | | |
| 32 | Anxiety | 2 | 0 | 0.13 | 0.00 | 0.00 | 1.00 |
| 33 | Aortic athero. | 2 | 0 | 0.19 | 0.00 | 0.00 | 1.00 |
| 34 | APD | 2 | 0 | 0.05 | 0.00 | 0.00 | 1.00 |
| 35 | CAD | 2 | 0 | 0.19 | 0.00 | 0.00 | 1.00 |
| 36 | Coronary rev. | 2 | 0 | 0.11 | 0.00 | 0.00 | 1.00 |
| 37 | Diabetes | 2 | 0 | 0.24 | 0.00 | 0.00 | 1.00 |
| 38 | Family history of CAD | 2 | 0 | 0.06 | 0.00 | 0.00 | 1.00 |
| 39 | Hypercholesteremia | 2 | 1 | 0.54 | 1.00 | 0.00 | 1.00 |
| 40 | Hypertension | 2 | 1 | 0.53 | 1.00 | 0.00 | 1.00 |
| 41 | Myocardial infarction | 2 | 0 | 0.12 | 0.00 | 0.00 | 1.00 |
| 42 | PAD | 2 | 0 | 0.04 | 0.00 | 0.00 | 1.00 |
| 43 | Pre-diabetes | 2 | 0 | 0.14 | 0.00 | 0.00 | 1.00 |
| 44 | Prior catheter | 2 | 0 | 0.02 | 0.00 | 0.00 | 1.00 |
| 45 | Prior CT | 2 | 0 | 0.00 | 0.00 | 0.00 | 1.00 |
| 46 | Prior echo | 2 | 0 | 0.00 | 0.00 | 0.00 | 1.00 |
| 47 | Prior MPI | 2 | 0 | 0.03 | 0.00 | 0.00 | 1.00 |
| 48 | Prior treadmill | 2 | 0 | 0.03 | 0.00 | 0.00 | 1.00 |
| 49 | Prior UTL | 2 | 0 | 0.08 | 0.00 | 0.00 | 1.00 |
| 50 | Smoking | 2 | 0 | 0.11 | 0.00 | 0.00 | 1.00 |
| 51 | Stroke | 2 | 0 | 0.09 | 0.00 | 0.00 | 1.00 |
| **Missingness indicators** | | | | | | | |
| 52 | Missing BMI | 2 | 0 | 0.02 | 0.00 | 0.00 | 1.00 |
| 53 | Missing Diaphoresis | 2 | 0 | 0.35 | 0.00 | 0.00 | 1.00 |
| 54 | Missing ECG | 2 | 0 | 0.02 | 0.00 | 0.00 | 1.00 |
| 55 | Missing Exertion | 2 | 1 | 0.56 | 1.00 | 0.00 | 1.00 |
| 56 | Missing GFR | 2 | 0 | 0.09 | 0.00 | 0.00 | 1.00 |

**Table S1.** Summary of predictors *(continued)*

| | Name | Unique values | Mode | Mean | Median | Minimum | Maximum |
|---|---|---|---|---|---|---|---|
| 57 | Missing HbA1c | 2 | 1 | 0.60 | 1.00 | 0.00 | 1.00 |
| 58 | Missing HDL | 2 | 0 | 0.43 | 0.00 | 0.00 | 1.00 |
| 59 | Missing LDL | 2 | 0 | 0.39 | 0.00 | 0.00 | 1.00 |
| 60 | Missing O2 Saturation | 2 | 0 | 0.00 | 0.00 | 0.00 | 1.00 |
| 61 | Missing Obesity | 2 | 0 | 0.01 | 0.00 | 0.00 | 1.00 |
| 62 | Missing Pain on Inspiration | 2 | 1 | 0.71 | 1.00 | 0.00 | 1.00 |
| 63 | Missing Pain on Palpation | 2 | 1 | 0.69 | 1.00 | 0.00 | 1.00 |
| 64 | Missing Pulse | 2 | 0 | 0.00 | 0.00 | 0.00 | 1.00 |
| 65 | Missing Radiating Pain | 2 | 0 | 0.40 | 0.00 | 0.00 | 1.00 |
| 66 | Missing Respiration | 2 | 0 | 0.00 | 0.00 | 0.00 | 1.00 |
| 67 | Missing SBP | 2 | 0 | 0.00 | 0.00 | 0.00 | 1.00 |
| 68 | Missing Sharp Pain | 2 | 1 | 0.76 | 1.00 | 0.00 | 1.00 |
| 69 | Missing Total Cholesterol | 2 | 0 | 0.43 | 0.00 | 0.00 | 1.00 |
| 70 | Missing Triglcyerides | 2 | 0 | 0.47 | 0.00 | 0.00 | 1.00 |
| 71 | Missing Troponin 3HV | 2 | 0 | 0.03 | 0.00 | 0.00 | 1.00 |

#### eTable 2. Cross-validated precision-recall AUC discrimination performance

| Learner | PR-AUC | Std. Err. | CI Lower | CI Upper |
|---|---|---|---|---|
| Marginal mean | 0.0188 | 0.0000 | 0.0187 | 0.0188 |
| Stratification (EDACS) | 0.0463 | 0.0015 | 0.0435 | 0.0492 |
| Stratification (ECG) | 0.0468 | 0.0009 | 0.0449 | 0.0486 |
| Decision tree (trop.) | 0.0508 | 0.0007 | 0.0494 | 0.0523 |
| Decision tree pruned | 0.0508 | 0.0007 | 0.0494 | 0.0523 |
| Decision tree (trop., ECG) | 0.0566 | 0.0007 | 0.0553 | 0.0579 |
| Decision tree (trop., ECG, scores) | 0.0585 | 0.0004 | 0.0578 | 0.0592 |
| Decision tree | 0.0587 | 0.0003 | 0.0580 | 0.0593 |
| Logistic reg. (trop.) | 0.0633 | 0.0013 | 0.0607 | 0.0659 |
| Stratification (trop.) | 0.0636 | 0.0014 | 0.0610 | 0.0663 |
| Stratification (HEART) | 0.0639 | 0.0012 | 0.0616 | 0.0662 |
| Logistic reg. (trop., ECG) | 0.0831 | 0.0029 | 0.0775 | 0.0888 |
| Stratification (trop., ECG, EDACS) | 0.0869 | 0.0020 | 0.0830 | 0.0909 |
| Stratification (trop., ECG) | 0.0890 | 0.0023 | 0.0845 | 0.0935 |
| Decision tree tuned | 0.0932 | 0.0050 | 0.0835 | 0.1030 |
| Stratification (trop., ECG, HEART) | 0.0982 | 0.0020 | 0.0943 | 0.1021 |
| Logistic reg. (trop., ECG, scores) | 0.1032 | 0.0038 | 0.0957 | 0.1106 |
| Linear reg. | 0.1135 | 0.0019 | 0.1099 | 0.1171 |
| Lasso | 0.1206 | 0.0031 | 0.1146 | 0.1266 |
| Logistic reg. | 0.1217 | 0.0035 | 0.1148 | 0.1286 |
| XGBoost | 0.1263 | 0.0051 | 0.1162 | 0.1363 |
| Splines | 0.1273 | 0.0034 | 0.1206 | 0.1340 |
| BART | 0.1356 | 0.0036 | 0.1284 | 0.1427 |
| XGBoost tuned | 0.1393 | 0.0029 | 0.1337 | 0.1449 |
| DiscreteSL | 0.1426 | 0.0049 | 0.1329 | 0.1522 |
| Random forest | 0.1472 | 0.0064 | 0.1347 | 0.1597 |
| Random forest tuned | 0.1486 | 0.0066 | 0.1356 | 0.1616 |
| SuperLearner | 0.1508 | 0.0049 | 0.1412 | 0.1603 |

**eTable 3. Cross-validated receiver operating characteristic AUC discrimination performance**

| Learner | ROC-AUC | Std. Err. | CI Lower | CI Upper |
|---|---|---|---|---|
| Marginal mean | 0.5000 | 0.0108 | 0.4789 | 0.5211 |
| Stratification (ECG) | 0.6870 | 0.0084 | 0.6705 | 0.7035 |
| Decision tree (trop.) | 0.6983 | 0.0106 | 0.6776 | 0.7190 |
| Decision tree pruned | 0.6983 | 0.0106 | 0.6776 | 0.7190 |
| Logistic reg. (trop.) | 0.7001 | 0.0084 | 0.6837 | 0.7166 |
| Stratification (trop.) | 0.7065 | 0.0084 | 0.6901 | 0.7230 |
| Stratification (EDACS) | 0.7339 | 0.0052 | 0.7237 | 0.7441 |
| Logistic reg. (trop., ECG) | 0.7611 | 0.0065 | 0.7484 | 0.7737 |
| Decision tree (trop., ECG) | 0.7635 | 0.0073 | 0.7492 | 0.7778 |
| Stratification (HEART) | 0.7647 | 0.0054 | 0.7541 | 0.7753 |
| Stratification (trop., ECG) | 0.7718 | 0.0066 | 0.7588 | 0.7849 |
| Stratification (trop., ECG, EDACS) | 0.7742 | 0.0058 | 0.7628 | 0.7855 |
| Decision tree (trop., ECG, scores) | 0.7827 | 0.0069 | 0.7691 | 0.7963 |
| Decision tree | 0.7835 | 0.0069 | 0.7699 | 0.7970 |
| Decision tree tuned | 0.8035 | 0.0054 | 0.7930 | 0.8140 |
| Logistic reg. (trop., ECG, scores) | 0.8122 | 0.0044 | 0.8035 | 0.8209 |
| Stratification (trop., ECG, HEART) | 0.8122 | 0.0048 | 0.8028 | 0.8217 |
| Linear reg. | 0.8406 | 0.0044 | 0.8320 | 0.8491 |
| Logistic reg. | 0.8447 | 0.0039 | 0.8371 | 0.8523 |
| Lasso | 0.8450 | 0.0038 | 0.8374 | 0.8525 |
| XGBoost | 0.8463 | 0.0039 | 0.8386 | 0.8539 |
| Splines | 0.8519 | 0.0038 | 0.8445 | 0.8593 |
| Random forest | 0.8610 | 0.0037 | 0.8537 | 0.8682 |
| XGBoost tuned | 0.8619 | 0.0036 | 0.8549 | 0.8690 |
| BART | 0.8628 | 0.0035 | 0.8559 | 0.8698 |
| DiscreteSL | 0.8630 | 0.0036 | 0.8560 | 0.8700 |
| Random forest tuned | 0.8647 | 0.0036 | 0.8577 | 0.8716 |
| SuperLearner | 0.8695 | 0.0034 | 0.8627 | 0.8763 |

#### eTable 4. Comparing missing value imputation using GLRM versus median/mode

| Variable | Missingness | Error GLRM | Error Median | Percent reduction |
|---|---|---|---|---|
| HbA1c | 59.8 | 0.088 | 1.628 | 94.6 |
| Triglycerides | 46.9 | 0.039 | 0.528 | 92.6 |
| HDL | 43.5 | 1.077 | 14.815 | 92.7 |
| Total Chol. | 42.7 | 7.961 | 45.762 | 82.6 |
| LDL | 38.8 | 4.158 | 37.291 | 88.8 |
| GFR | 8.8 | 0.286 | 11.699 | 97.6 |
| Trop. 3HV | 2.7 | 0.001 | 0.008 | 90.2 |
| ECG | 1.8 | 0.000 | 0.729 | 100.0 |
| BMI | 1.6 | 0.645 | 7.508 | 91.4 |
| Obese | 1.4 | 0.769 | 0.640 | -20.1 |
| Respiration | 0.4 | 0.020 | 2.909 | 99.3 |
| O2 Saturation | 0.3 | 0.020 | 2.515 | 99.2 |
| Pulse | 0.2 | 0.680 | 18.446 | 96.3 |
| Pulse Peak | 0.2 | 0.703 | 18.553 | 96.2 |
| SBP | 0.1 | 0.586 | 22.843 | 97.4 |
| Lowest SBP | 0.1 | 0.458 | 18.673 | 97.5 |

#### eTable 5. Cross-validated index of prediction accuracy for each learner and the ensemble

**Table S2.** Cross-validated index of prediction accuracy for each learner and the ensemble

| Learner | IPA | Std. Err. | CI Lower | CI Upper |
|---|---|---|---|---|
| Marginal mean | 0.0000 | 0.0000 | 0.0000 | 0.0000 |
| Stratification (EDACS) | 0.0144 | 0.0008 | 0.0128 | 0.0159 |
| Stratification (ECG) | 0.0156 | 0.0007 | 0.0142 | 0.0170 |
| Logistic reg. (trop.) | 0.0190 | 0.0011 | 0.0168 | 0.0212 |
| Decision tree (trop.) | 0.0235 | 0.0007 | 0.0220 | 0.0250 |
| Decision tree pruned | 0.0235 | 0.0007 | 0.0220 | 0.0250 |
| Stratification (HEART) | 0.0245 | 0.0007 | 0.0231 | 0.0259 |
| Stratification (trop., ECG, EDACS) | 0.0257 | 0.0026 | 0.0206 | 0.0309 |
| Decision tree (trop., ECG) | 0.0271 | 0.0007 | 0.0257 | 0.0284 |
| Stratification (trop.) | 0.0274 | 0.0007 | 0.0261 | 0.0287 |
| Decision tree (trop., ECG, scores) | 0.0282 | 0.0001 | 0.0279 | 0.0285 |
| Decision tree | 0.0283 | 0.0002 | 0.0279 | 0.0288 |
| Logistic reg. (trop., ECG) | 0.0318 | 0.0021 | 0.0277 | 0.0359 |
| Stratification (trop., ECG) | 0.0389 | 0.0012 | 0.0364 | 0.0413 |
| Decision tree tuned | 0.0395 | 0.0018 | 0.0359 | 0.0430 |
| Stratification (trop., ECG, HEART) | 0.0438 | 0.0016 | 0.0407 | 0.0470 |
| Logistic reg. (trop., ECG, scores) | 0.0453 | 0.0023 | 0.0408 | 0.0497 |
| XGBoost | 0.0459 | 0.0046 | 0.0370 | 0.0549 |
| Linear reg. | 0.0465 | 0.0002 | 0.0462 | 0.0469 |
| Logistic reg. | 0.0556 | 0.0018 | 0.0521 | 0.0590 |
| Lasso | 0.0560 | 0.0016 | 0.0528 | 0.0591 |
| Splines | 0.0584 | 0.0018 | 0.0549 | 0.0618 |
| BART | 0.0666 | 0.0024 | 0.0618 | 0.0714 |
| XGBoost tuned | 0.0685 | 0.0020 | 0.0645 | 0.0725 |
| DiscreteSL | 0.0696 | 0.0025 | 0.0647 | 0.0745 |
| Random forest | 0.0705 | 0.0035 | 0.0637 | 0.0774 |
| Random forest tuned | 0.0708 | 0.0029 | 0.0652 | 0.0764 |
| SuperLearner | 0.0757 | 0.0030 | 0.0698 | 0.0815 |

#### eTable 6. Cross-validated Brier score for each learner and the ensemble

**Table S3.** Cross-validated Brier score for each learner and the ensemble

| Learner | Brier score | Std. Err. | CI Lower | CI Upper |
|---|---|---|---|---|
| Marginal mean | 0.01840 | 1e-05 | 0.01839 | 0.01842 |
| Stratification (EDACS) | 0.01814 | 2e-05 | 0.01810 | 0.01818 |
| Stratification (ECG) | 0.01812 | 1e-05 | 0.01810 | 0.01813 |
| Logistic reg. (trop.) | 0.01805 | 2e-05 | 0.01802 | 0.01809 |
| Decision tree (trop.) | 0.01797 | 2e-05 | 0.01793 | 0.01801 |
| Decision tree pruned | 0.01797 | 2e-05 | 0.01793 | 0.01801 |
| Stratification (HEART) | 0.01795 | 1e-05 | 0.01793 | 0.01798 |
| Stratification (trop., ECG, EDACS) | 0.01793 | 5e-05 | 0.01782 | 0.01804 |
| Decision tree (trop., ECG) | 0.01791 | 1e-05 | 0.01788 | 0.01793 |
| Stratification (trop.) | 0.01790 | 1e-05 | 0.01787 | 0.01793 |
| Decision tree (trop., ECG, scores) | 0.01788 | 1e-05 | 0.01786 | 0.01790 |
| Decision tree | 0.01788 | 1e-05 | 0.01786 | 0.01790 |
| Logistic reg. (trop., ECG) | 0.01782 | 4e-05 | 0.01775 | 0.01789 |
| Stratification (trop., ECG) | 0.01769 | 2e-05 | 0.01765 | 0.01773 |
| Decision tree tuned | 0.01768 | 3e-05 | 0.01762 | 0.01774 |
| Stratification (trop., ECG, HEART) | 0.01760 | 3e-05 | 0.01754 | 0.01765 |
| Logistic reg. (trop., ECG, scores) | 0.01757 | 4e-05 | 0.01749 | 0.01765 |
| XGBoost | 0.01756 | 8e-05 | 0.01739 | 0.01772 |
| Linear reg. | 0.01755 | 1e-05 | 0.01753 | 0.01756 |
| Logistic reg. | 0.01738 | 3e-05 | 0.01732 | 0.01744 |
| Lasso | 0.01737 | 3e-05 | 0.01732 | 0.01742 |
| Splines | 0.01733 | 3e-05 | 0.01728 | 0.01738 |
| BART | 0.01718 | 4e-05 | 0.01709 | 0.01727 |
| XGBoost tuned | 0.01714 | 4e-05 | 0.01707 | 0.01721 |
| DiscreteSL | 0.01712 | 5e-05 | 0.01703 | 0.01722 |
| Random forest | 0.01711 | 7e-05 | 0.01698 | 0.01723 |
| Random forest tuned | 0.01710 | 5e-05 | 0.01700 | 0.01720 |
| SuperLearner | 0.01701 | 5e-05 | 0.01690 | 0.01712 |

##### eTable 7. Grouped calibration table

| Decile | N | Predicted risk | Observed risk | Predicted / Actual | Predicted - Actual |
|---|---|---|---|---|---|
| 1 | 11,672 | 0.045% | 0.026% | 1.74 | 0.019% |
| 2 | 11,671 | 0.100% | 0.043% | 2.33 | 0.057% |
| 3 | 11,671 | 0.177% | 0.129% | 1.38 | 0.049% |
| 4 | 11,671 | 0.297% | 0.231% | 1.29 | 0.066% |
| 5 | 11,671 | 0.489% | 0.471% | 1.04 | 0.018% |
| 6 | 11,671 | 0.790% | 0.651% | 1.21 | 0.139% |
| 7 | 11,671 | 1.272% | 1.122% | 1.13 | 0.149% |
| 8 | 11,671 | 2.089% | 1.791% | 1.17 | 0.298% |
| 9 | 11,671 | 3.746% | 3.624% | 1.03 | 0.122% |
| 10 | 11,671 | 10.519% | 10.667% | 0.99 | -0.148% |

##### eTable 8. Net unnecessary interventions avoided per 100 patients at 0.5% threshold

| Estimator | True positive rate | False positive rate | Net benefit | Net interventions avoided |
|---|---|---|---|---|
| Treat All | 1.88% | 98.12% | 0.0138 | 0.0 |
| Decision tree | 1.58% | 37.44% | 0.0139 | 1.7 |
| EDACS | 1.80% | 80.73% | 0.0140 | 2.9 |
| HEART | 1.81% | 71.94% | 0.0145 | 13.9 |
| Logistic reg. | 1.80% | 58.25% | 0.0151 | 24.9 |
| Random forest | 1.78% | 51.15% | 0.0153 | 28.7 |
| SuperLearner | 1.81% | 52.56% | 0.0154 | 31.9 |

#### References


Austin, Peter C (2007). "A comparison of regression trees, logistic regression, generalized additive models, and multivariate adaptive regression splines for predicting AMI mortality". In: *Statistics in medicine* 26.15, pp. 2937–2957.

Benkeser, David, Maya Petersen, and Mark J van der Laan (2019). "Improved small-sample estimation of nonlinear cross-validated prediction metrics". In: *Journal of the American Statistical Association*, pp. 1–16.

Breiman, Leo (2001). "Random forests". In: *Machine learning* 45.1, pp. 5–32.

Brooks, Jordan, Mark J van der Laan, and Alan S Go (2012). "Targeted maximum likelihood estimation for prediction calibration". In: *The international journal of biostatistics* 8.1.

Effrosynidis, Dimitrios and Avi Arampatzis (2021). "An evaluation of feature selection methods for environmental data". In: *Ecological Informatics* 61, p. 101224.

Hastie, Trevor, Robert Tibshirani, and Ryan J Tibshirani (2017). "Extended comparisons of best subset selection, forward stepwise selection, and the lasso". In: *arXiv preprint arXiv:1707.08692*.

Hubbard, Alan E, Chris J Kennedy, and Mark J van der Laan (2018). "Data-Adaptive Target Parameters". In: *Targeted Learning in Data Science*. Springer, pp. 125–142.

Jaeger, Byron C, Nicholas J Tierney, and Noah R Simon (2020). "When to Impute? Imputation before and during cross-validation". In: *arXiv preprint arXiv:2010.00718*.

Li, Lisha, Kevin Jamieson, Giulia DeSalvo, Afshin Rostamizadeh, and Ameet Talwalkar (2017). "Hyperband: A novel bandit-based approach to hyperparameter optimization". In: *The Journal of Machine Learning Research* 18.1, pp. 6765–6816.

Molnar, Christoph (2020). *Interpretable Machine Learning*. Lulu. com.

Probst, Philipp and Anne-Laure Boulesteix (2017). "To tune or not to tune the number of trees in random forest". In: *The Journal of Machine Learning Research* 18.1, pp. 6673–6690.

Roberts, David R, Volker Bahn, Simone Ciuti, Mark S Boyce, Jane Elith, Gurutzeta Guillera-Arroita, Severin Hauenstein, José J Lahoz-Monfort, Boris Schröder, Wilfried Thuiller, et al. (2017). "Cross-validation strategies for data with temporal, spatial, hierarchical, or phylogenetic structure". In: *Ecography* 40.8, pp. 913–929.

Sisk, Rose, Lijing Lin, Matthew Sperrin, Jessica K Barrett, Brian Tom, Karla Diaz-Ordaz, Niels Peek, and Glen P Martin (2021). "Informative presence and observation in routine health data: A review of methodology for clinical risk prediction". In: *Journal of the American Medical Informatics Association* 28.1, pp. 155–166.

Sperrin, Matthew, Glen P Martin, Rose Sisk, and Niels Peek (2020). "Missing data should be handled differently for prediction than for description or causal explanation". In: *Journal of Clinical Epidemiology* 125, pp. 183–187.

Strobl, Carolin, Anne-Laure Boulesteix, Achim Zeileis, and Torsten Hothorn (2007). "Bias in random forest variable importance measures: Illustrations, sources and a solution". In: *BMC bioinformatics* 8.1, pp. 1–21.

Williamson, Brian D, Peter B Gilbert, Noah Simon, and Marco Carone (2017). "Nonparametric variable importance assessment using machine learning techniques". In:
